# Figurative Drawing Ability is Associated with Advanced Compositional Language Comprehension in Neurodevelopmental Disorders

**DOI:** 10.1101/2024.07.26.24310995

**Authors:** Elliot Murphy, Rohan Venkatesh, Edward Khokhlovich, Andrey Vyshedskiy

## Abstract

In research on human language evolution, drawing is often treated as a unitary marker of symbolic cognition, yet non-figurative and figurative drawing may depend on partially distinct cognitive capacities. We examined the relationship between caregiver-reported drawing abilities and language phenotypes in 77,616 children and adolescents with neurodevelopmental disorders. Participants were assessed using a 133-item caregiver questionnaire that included receptive-language, expressive-language, and drawing items. Building on prior work identifying three clusters of co-occurring receptive-language abilities (Command, Modifier, and Syntactic mechanisms) and three clusters expressive-language abilities (Single-Word, Single-Sentence, and Multi-Sentence production), we used unsupervised hierarchical clustering and principal component analysis to determine where drawing abilities fall within this broader cognitive-developmental space. Non-figurative drawing clustered most closely with the Modifier mechanism, whereas two figurative drawing items (*drawing a variety of recognizable images* and *drawing a novel image from verbal description*) clustered with the Syntactic mechanism. These associations were broadly stable across age, sex, severity, and native-language subgroups. The findings suggest that (1) drawing proficiency relates more strongly to language comprehension than to speech, and (2) drawing abilities are not uniformly associated with comprehension development: broad non-figurative graphic activity aligns with modifier-level comprehension, whereas figurative drawing is more closely linked to compositional syntactic comprehension.

## Introduction

Drawing has long been used as a window into symbolic abilities, both for monitoring child development and for diagnosing dementia ^1–3^. The study of human evolution is no exception, with symbolic capacities frequently being inferred from evidence of drawing. Discoveries of artistic artifacts in Eurasia that predate the migration of *Homo sapiens* have sparked significant excitement. Pre-*Homo sapiens* artistic artifacts include the use of pigments ^4^, perforated shells ^4–8^, line marks on stones and shells ^9,10^, geometrical figures ^11^, and hand stencils painted on cave walls ^12–18^. While these artifacts may be symbolic, they are non-figurative – they do not represent clear images of humans, animals, or other recognizable objects.

Currently known early figurative traditions are substantially later than many non-figurative marks and ornaments, with prominent examples from Island Southeast Asia and Europe dating to roughly 44-38kya. The earliest examples of figurative art include a hunting scene depicting part-human, part-animal figures from the limestone cave of Leang Bulu’ Sipong 4 (Sulawesi, Indonesia), dated to 44kya ^19^; a painting of an animal at a limestone cave in Indonesian Borneo, dated to 40kya ^20^; a drawing of a “deer-pig" from the Indonesian island of Sulawesi, also dated to 40kya ^21^; an engraving of an extinct wild cow (aurochs) from the Dordogne region of France, dated to 38kya ^22^; and the lion-man sculpture excavated from the caves of Lone Valley in Germany, dated to 39kya ^23^.

Does the transition from non-figurative to figurative traditions reflect a fundamental shift in symbolic cognition? One approach to tentatively examining the relationship between artistic capacities and cognition is the study of modern individuals with neurodevelopmental disorders. Many such individuals harbor deleterious mutations in genes implicated in broad aspects of intellectual functions and speech, including *FOXP2* ^24–26^, *SRGAP2A* ^27^, and *CNTNAP2* ^28^. Some neurodevelopmental disorders affect pathways relevant to language, cognition, motor control, and symbolic behavior. While contemporary clinical populations cannot be treated as direct models of ancestral cognition, they can reveal dissociations among abilities that are difficult to observe in typically developing children.

The motivation for this study was two-fold. First, we had access to the largest database of over 100,000 individuals exhibiting a range of symbolic abilities, with parent-reported figurative and non-figurative drawing skills. Second, previous work has demonstrated distinct clustering of receptive and expressive language abilities ^29–32^. Access to this data enabled the present investigation of associations between drawing skills and language phenotypes. Specifically, we examine how drawing abilities co-occur with different language phenotypes. If drawing abilities were causally independent of language phenotypes – like unrelated traits such as hyperactivity – they would be expected to occur with equal frequency across language levels and to form a cluster separate from the receptive and expressive language clusters. Conversely, an association between a drawing ability and a specific language phenotype would tentatively point to possible maturational links between symbolic drawing skills and language proficiency (though these need not necessarily be causally connected).

A central challenge in debates about art, symbolism, and language evolution is that “symbolic cognition” is often treated as a single capacity. Yet the production of a hand stencil, a geometric engraving, a recognizable animal, and a novel imagined creature may place very different demands on perception, motor planning, visual memory, mental imagery, and relational/combinatorial cognition. Non-figurative marks may require intentional graphic production without requiring the construction of a mental representation of a recognizable object. Figurative drawing, by contrast, requires the selection and organization of visual features into a configuration that can be recognized by another observer as a particular kind of object, animal, person, or imagined entity. Drawing a novel figure from verbal description adds a further demand: the individual must transform linguistic input into an internally generated visual representation that is not simply copied from memory.

For this reason, the present study does not ask whether drawing, in general, is related to language. Rather, it asks whether different forms of drawing map onto different language-comprehension phenotypes. This distinction is important because a broad association between drawing and language could simply reflect general developmental level. By contrast, a selective association between non-figurative drawing and one language cluster, and figurative drawing and another, would suggest a more structured relationship between graphic production and specific forms of linguistic-combinatorial cognition.

## Methods

### Study Participants

Participants were children and adolescents using a language therapy app that was made freely available at all major app stores in September 2015 ^33–37^. The app provides various structured language comprehension therapy exercises and is primarily used by caregivers of children with language impairments. Most of the caregivers are presumed to be parents. Once the app was downloaded, caregivers were asked to register and to provide demographic details, including the child’s diagnosis and age. Caregivers completed a 133-item questionnaire approximately every three months: 77-item Autism Treatment Evaluation Checklist (ATEC) ^38^ (Supplementary Tables 1-4); 20-item Mental Synthesis Evaluation Checklist (MSEC) ^39^ (Supplementary Table 5); 10-item screen time checklist ^40^; 25-item diet checklist ^41^; and 1-item parent education survey.

All 26 language-related items (15 receptive, Table 1, and 11 expressive, Table 2) available in the 133-item questionnaire were included in the cluster analysis as in previously published articles ^29–32^. Response options were: very true (0 points), somewhat true (1 point), and not true (2 points).

**Table 1.** Three language comprehension mechanisms—Syntactic, Modifier, and Command—have been identified in previous studies. ^29–31^. When one mechanism is acquired, the entire range of associated comprehension abilities is also gained. The Command-level abilities (Items 1 to 4) are acquired first. The Modifier-level abilities (Items 5 to 8) are attained next. The Syntactic-level abilities (Items 9 to 15) are acquired last. The language comprehension items are presented exactly as surveyed with parents in both this and earlier studies. Response options were: very true, somewhat true, and not true. Items 1 to 3 were assessed as part of the Expressive Language ATEC ^38^ subscale 1; the rest of items were a part of the MSEC subscale ^39^.

|  |  | Language comprehension items (verbatim) | Abbreviations used in dendrograms |
| --- | --- | --- | --- |
| Command Mechanism | 1 | Knows own name | Knows Name |
|  | 2 | Responds to 'No' or 'Stop' | No and Stop |
|  | 3 | Responds to praise | Resp. to Praise |
|  | 4 | Can follow some commands | Commands |
| Modifier Mechanism | 5 | Understands some simple modifiers (i.e., green apple vs. red apple or big apple vs. small apple) | Color or Size / Modifiers |
|  | 6 | Understands several modifiers in a sentence (i.e., small green apple) | Two Modifiers |
|  | 7 | Understands size (can select the largest/smallest object out of a collection of objects) | Size Superlatives |
|  | 8 | Understands NUMBERS (i.e., two apples vs. three apples) | Numbers |
| Syntactic Mechanism | 9 | Understands spatial prepositions (i.e., put the apple ON TOP of the box vs. INSIDE the box vs. BEHIND the box) | Sp. Prepositions |
|  | 10 | Understands verb tenses (i.e., I will eat an apple vs. I ate an apple) | Verb Tenses |
|  | 11 | Understands simple stories that are read aloud | Simple Stories |
|  | 12 | Understands elaborate fairytales that are read aloud (i.e., stories describing FANTASY creatures) | Elab. Fairytales |
|  | 13 | Understands possessive pronouns (i.e., your apple vs. her apple) | Poss. Pronouns |
|  | 14 | Understands the change in meaning when the order of words is changed (i.e., understands the difference between 'a cat ate a mouse' vs. 'a mouse ate a cat') | Flexible Syntax |
|  | 15 | Understands explanations about people, objects or situations beyond the immediate surroundings (e.g., "Mom is walking the dog," "The snow has turned to water"). | Explanations |

**Table 2.** Eleven expressive language items as they were posed to parents. Response options were: very true (0 points), somewhat true (1 point), and not true (2 points). All items were a part of the expressive language ATEC subscale 1. Single-Word, Single-Sentence, and Multi-Sentence clusters have been identified in previous work ^32^.

|  |  | Expressive language items (verbatim) | Abbreviations used in dendrograms |
| --- | --- | --- | --- |
| Single-Word | 1 | [My child] Can use one word at a time (No!, Eat, Water, etc.) | Uses 1 word |
|  | 2 | Knows 10 or more words | Knows 10+ words |
| Single-Sentence | 3 | Can use 2 words at a time (Don't want, Go home) | Uses 2 words |
|  | 4 | Can use 3 words at a time (Want more milk) | Uses 3 words |
|  | 5 | Can use sentences with 4 or more words | Uses 4+ words |
|  | 6 | Explains what he/she wants | Explains what wants |
|  | 7 | Speech tends to be meaningful/relevant | Speech meaningful |
| Multi-Sentence | 8 | Carries on fairly good conversation | Good conversation |
|  | 9 | Has normal ability to communicate for his/her age | Normal communic. |
|  | 10 | Asks meaningful questions | Asks questions |
|  | 11 | Often uses several successive sentences | Uses several sentences |

The inclusion criteria for this study remained consistent with those of previous studies ^29–32^: absence of seizures (which commonly result in intermittent, unstable language deficits ^42,43^), absence of serious and moderate sleep problems (which are also associated with intermittent, unstable language deficits ^44^), age range of 4 to 22 years (the lower age cutoff was chosen to ensure that participants were exposed to complete sets of items listed in Table 1 ^45^, whereas the upper age threshold was constrained by the scarcity of older participants).

Table 3 reports participants’ diagnoses as communicated by caregivers. Autism level (mild/Level 1, moderate/Level 2, or severe/Level 3) was reported by caregivers. Pervasive Developmental Disorder and Asperger Syndrome were combined with mild autism for analyses, as recommended by DSM-5 ^46^. A good reliability of such parent-reported diagnosis has been previously demonstrated ^47^.

**Table 3.** Participants’ diagnoses.

|  | **Number of**<br>**Participants** | **Percent of**<br>**Total** | **Age, Mean(SD)** | **Percent Males** |
| --- | --- | --- | --- | --- |
| **Mild Autism Spectrum Disorder (ASD)** | 31072 | 40.0 | 6.3(2.4) | 76.8 |
| **Moderate ASD** | 19844 | 25.6 | 6.9(2.8) | 80 |
| **Severe ASD** | 11498 | 14.8 | 7.6(3.3) | 80.6 |
| **Specific Language Impairment** | 4928 | 6.3 | 6.0(2.2) | 69.1 |
| **Mild Language Delay** | 4024 | 5.2 | 5.5(1.7) | 67.4 |
| **ADHD** | 1649 | 2.1 | 6.4(2.2) | 72.8 |
| **Down Syndrome** | 1450 | 1.9 | 8.5(3.5) | 60.4 |
| **Other Genetic Disorder** | 1120 | 1.4 | 8.0(3.5) | 57.7 |
| **Social Communication Disorder** | 805 | 1.0 | 6.3(2.3) | 69.1 |
| **Sensory Processing Disorder** | 754 | 1.0 | 6.8(2.7) | 70.2 |
| **Apraxia** | 472 | 0.6 | 6.8(2.9) | 64.6 |
| **Total** | 77,616 | 100.0 | 6.6(2.7) | 76.3 |

When caregivers completed several evaluations, the last evaluation was used for analysis. Thus, the study included a total of 77,616 participants, the average age was 6.6±2.7 years (range of 4 to 22 years), 75.7% participants were males (Table 3). The education level of participants’ parents was the following: 90.8% with at least a high school diploma, 68.7% with at least college education, 35.9% with at least a master’s qualification, and 5.7% with a doctorate degree.

The study was conducted in accordance with the Declaration of Helsinki ^48^. Informed consent was obtained from the caregivers of all participants. The study protocol was approved by the Biomedical Research Alliance of New York (BRANY) LLC Institutional Review Board.

### Drawing assessments

The evaluation included three questions about participants’ drawing abilities. One item assessed general drawing ability, including non-figurative drawing: *“[My child] does drawing, coloring, art*”; and two items assessed figurative drawing ability: *“[My child] draws a VARIETY of RECOGNIZABLE images (objects, people, animals, etc.)*” and *“[My child] can draw a NOVEL image following YOUR description (e.g. a three-headed horse)*”. Response options were “very true”, “somewhat true”, and “not true”.

These three drawing items should be interpreted as broad caregiver-reported indices rather than formal psychometric measures of drawing ability. The non-figurative item captures participation in drawing, coloring, or art activities and therefore may include scribbling, coloring, tracing, or other graphic activity that does not necessarily produce recognizable figures. The two figurative items were designed to capture more specific representational abilities: the production of a variety of recognizable images and the production of a novel image from verbal description. The latter item places additional demands on verbal comprehension, mental imagery, and the transformation of a linguistic description into a visual plan.

### Statistical analysis

Unsupervised Hierarchical Cluster Analysis (UHCA) is a robust method for detecting patterns of co-occurring abilities within noisy data. UHCA is an exploratory, data-driven approach, although results can depend on choices of distance metric, linkage method, and item scaling. UHCA was performed using Ward’s agglomeration method with a Euclidean distance metric.

An additional statistical approach, Principal Component Analysis (PCA), was used to confirm the UHCA findings. PCA employs a fundamentally different mathematical framework for dimensionality reduction, transforming high-dimensional, noisy measures into a compact set of orthogonal components that capture the dominant sources of variance in the data. This data-driven projection spatially organizes abilities that co-occur more frequently closer together in a two-dimensional component space, providing an independent convergence test for the clustering structure. Analyses were performed in R^49^. Code and data are publicly available (https://doi.org/10.17605/OSF.IO/3ZYJ8).

Our large dataset using a 3-point Likert scale (0–2) is well suited for UHCA and PCA because it provides ordered categorical data that can be treated as approximately continuous ^50,51^. First, while mathematical distortions in ordinal data can be exacerbated by severe distribution skewness, a 3-point scale strictly constrains the metric space. Because the data are bounded within a narrow (0–2) grid, there is no possibility for extreme outlier values to disproportionately warp the Euclidean distance calculations ^52^. Second, whereas small datasets can suffer from fragmented or unstable clustering due to violations of the equal-intervals assumption, large datasets benefit from asymptotic smoothing ^50,53^. In large datasets, macro-level aggregate patterns dominate individual-level discrete variability, allowing this approach to achieve an optimal balance between geometric variance and structural stability.

Because UHCA and PCA are descriptive multivariate techniques, the resulting clusters should not be interpreted as evidence of causal relations among abilities. Rather, they identify patterns of co-occurrence among questionnaire items. We therefore use clustering to test whether drawing items align more closely with particular receptive or expressive language phenotypes than would be expected if drawing abilities were unrelated to language structure.

## Results

### Clustering analysis of language comprehension abilities

Fifteen language comprehension abilities were assessed by caregivers (Table 1). To examine patterns of co-occurrence among these abilities, we applied unsupervised hierarchical cluster analysis (UHCA), a data-driven method that produces tree-like diagrams, called dendrograms, which visually represent hierarchical relationships between clusters of items. Abilities that frequently co-occur are automatically positioned closer together, while those that co-occur less often appear farther apart.

The UHCA-derived dendrogram (Figure 1A) replicated the hierarchical cluster structure reported in our prior work ^29–31^. The first cluster included “*[My child] knows his/her name*”, “*[My child] responds to ‘No’ or ‘Stop’”, “[My child] responds to praise”,* and “*[My child] follows some commands*” (items 1 to 4 in Table 1), and was previously termed the Command Mechanism. The second cluster included “*[My child] understands color and size modifiers”,* “*[My child] understands several modifiers in a sentence”,* “*[My child] understands size superlatives”,* and “*[My child] understands numbers”* (items 5 to 8 in Table 1), and was designated as the Modifier Mechanism in our prior work. The third cluster included “*[My child] understands spatial prepositions”,* “*[My child] understands verb tenses”,* “*[My child] understands flexible syntax”,* “*[My child] understands possessive pronouns”,* “*[My child] understands explanations about people and situations”,* “*[My child] understands simple stories”,* and “*[My child] understands elaborate fairytales”* (items 9 to 15 in Table 1), and was previously termed the Syntactic Mechanism.

**Figure 1.**
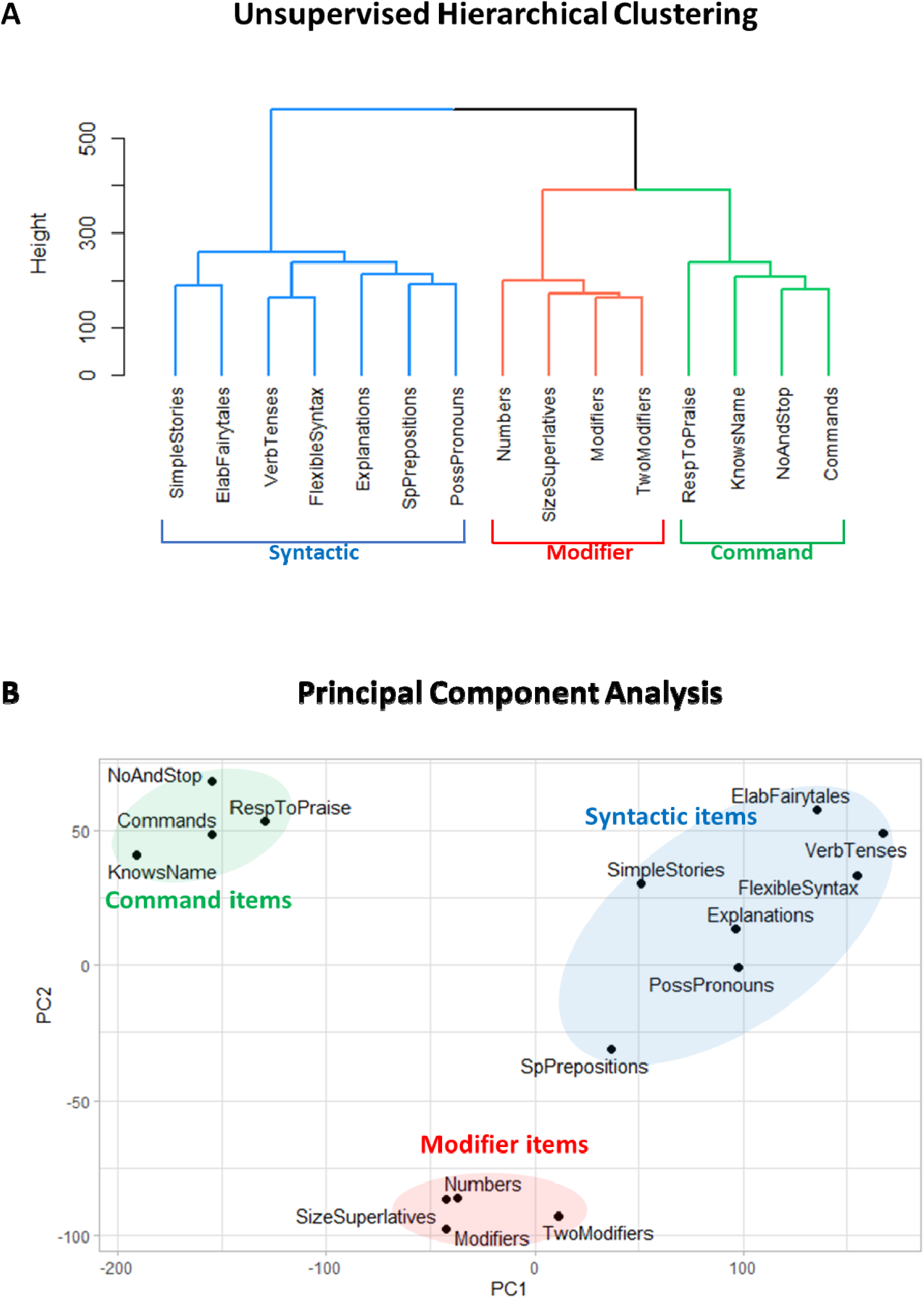
Clustering analysis of 15 language comprehension items. (A) The dendrogram representing the hierarchical clustering of language comprehension abilities. (B) Principal component analysis of language comprehension abilities shows clear separation between Command, Modifier, and Syntactic items. Principal component 1 accounts for 46.1% of the variance in the data. Principal component 2 accounts for 11.2% of the variance in the data.

Principal component analysis (PCA) (Figure 1B) also showed a clear separation between the Command, Modifier, and Syntactic clusters. Each cluster occupied a unique region in the principal component space, with no overlap observed. This separation suggests that the underlying structure of the data effectively captures the functional differences among the clusters, providing strong evidence for their distinct classification.

Our previous analysis using both UHCA and PCA demonstrated that the Command, Modifier, and Syntactic clusters were highly stable across multiple evaluation methods (Ward, Average, Complete, McQuitty) ^29,30^, age groups (4 to 6 years, 6 to 12 years, 12 to 22 years) ^29,30^, time points (first vs. last evaluation) ^29,30^, genders ^29^, spoken-languages (English, Spanish, Portuguese, Italian, Russian, Chinese, French, German, or Korean) ^31^, diagnostic categories ^29,31^, and levels of parental education ^29^.

The reasons for the emergence of three distinct clusters of co-occurring abilities become clear when viewed through the lens of developmental dynamics. Some individuals with language impairments plateau at the Command level, others at the Modifier level, while only a subset progress to the Syntactic level ^54^. Individuals who plateau at the Command level typically acquire the full set of Command abilities; those who plateau at the Modifier level acquire both Command and Modifier abilities; and only individuals who reach the Syntactic level go on to acquire the full range of syntactic abilities. This developmental stratification explains the observed clustering pattern: the four Command abilities (items 1–4 in Table 1) reliably co-occur and are grouped by UHCA and PCA into the Command cluster; the four Modifier skills (items 5–8) co-occur and form the Modifier cluster; and the seven Syntactic abilities (items 9–15) co-occur and constitute the Syntactic cluster.

As a control, we calculated UHCA and PCA of the 15 language comprehension abilities along with the *hyperactivity* (Supplementary Figure 1) items. Since this item is not related to language, it would be expected to form a separate cluster. As anticipated, both UHCA and PCA grouped *hyperactivity* into its own cluster at a significant distance from the three comprehension clusters, indicating random co-occurrence with the language abilities, thus validating the effectiveness of both clustering techniques.

### Clustering analysis of expressive language abilities

Eleven expressive language abilities were assessed by caregivers (Table 2). The dendrogram generated by the UHCA of these abilities (Figure 2A) replicated the hierarchical cluster structure reported in our prior work ^32^. Three distinct clusters emerged. The first cluster, previously termed Single-Word, comprised the abilities “*[My child] can use one word at a time*” and “*[My child] knows 10 or more words*” (Table 2, items 1 and 2). The second cluster, termed Single-Sentence, included the abilities “*[My child] can use 2 words at a time*”, “*[My child] can use three words at a time*”, “*[My child] can use sentences with 4 or more words*”, “*[My child] explains what they want*”, and “*[My child] speech tends to be meaningful/relevant*” (items 3 to 7). The third cluster, termed Multi-Sentence, encompasses the abilities “*[My child] carries on fairly good conversation*”, “*[My child] has normal ability to communicate for their age*”*, “[My child] asks meaningful questions*”, and “*[My child] often uses several successive sentences”* (items 8 to 11). PCA (Figure 2B) also showed a clear separation between the Single-Word, Single-Sentence, and Multi-Sentence clusters. Previous analyses explored the stability of the expressive language clusters, which we refer the interested reader to ^32^.

**Figure 2.**
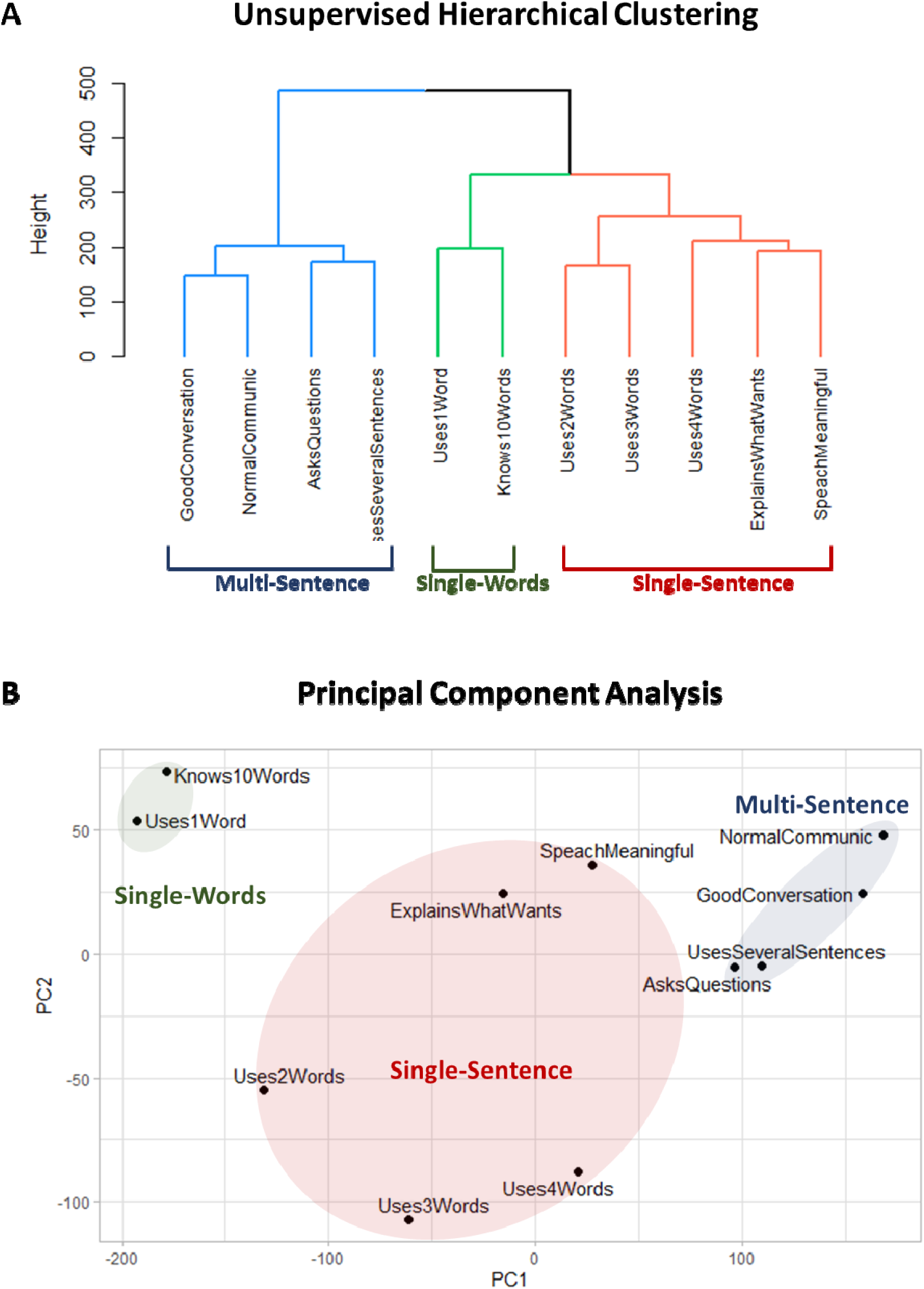
Clustering analysis of 11 expressive language items. (A) The dendrogram representing the unsupervised hierarchical clustering analysis of expressive language abilities. (B) Principal component analysis of expressive language abilities shows a clear separation between Single-Word, Single-Sentence, and Multi-Sentence items. Principal component 1 accounts for 51.2% of the variance in the data. Principal component 2 accounts for 10.9% of the variance in the data.

The reasons for the emergence of the three expressive language clusters mirror those underlying the three comprehension clusters. Specifically, some children plateau at the Single-Word level, others at the Single-Sentence level, while the majority progress to the Multi-Sentence level ^55^. Consequently, clustering analysis of large cohorts of children with language impairments consistently identify these developmental plateaus as sets of co-occurring abilities, reflecting the natural grouping of linguistic competencies.

As a control, we calculated UHCA and PCA of the 11 expressive language abilities along with the *hyperactivity* (Supplementary Figure 2) item. This item is not related to speech and was therefore expected to form a separate cluster. As anticipated, both UHCA and PCA grouped *hyperactivity* into its own cluster at a significant distance from the three expressive clusters, validating the effectiveness of both clustering techniques.

### Clustering analysis of language comprehension abilities together with expressive language abilities

Figure 3A depicts the UHCA dendrogram of the 15 language comprehension items (Table 1) together with the 11 expressive language items (Table 2). As in previous work ^32^, both the UHCA (Figure 3A) and the PCA (Figure 3B) show six clusters of items. These clusters are identical to those detected by separate clustering of receptive (Figure 1) and expressive (Figure 2) items: three language comprehension clusters (Command, Modifier, and Syntactic) and three expressive language clusters (Single-Word, Single-Sentence, and Multi-Sentence).

**Figure 3.**
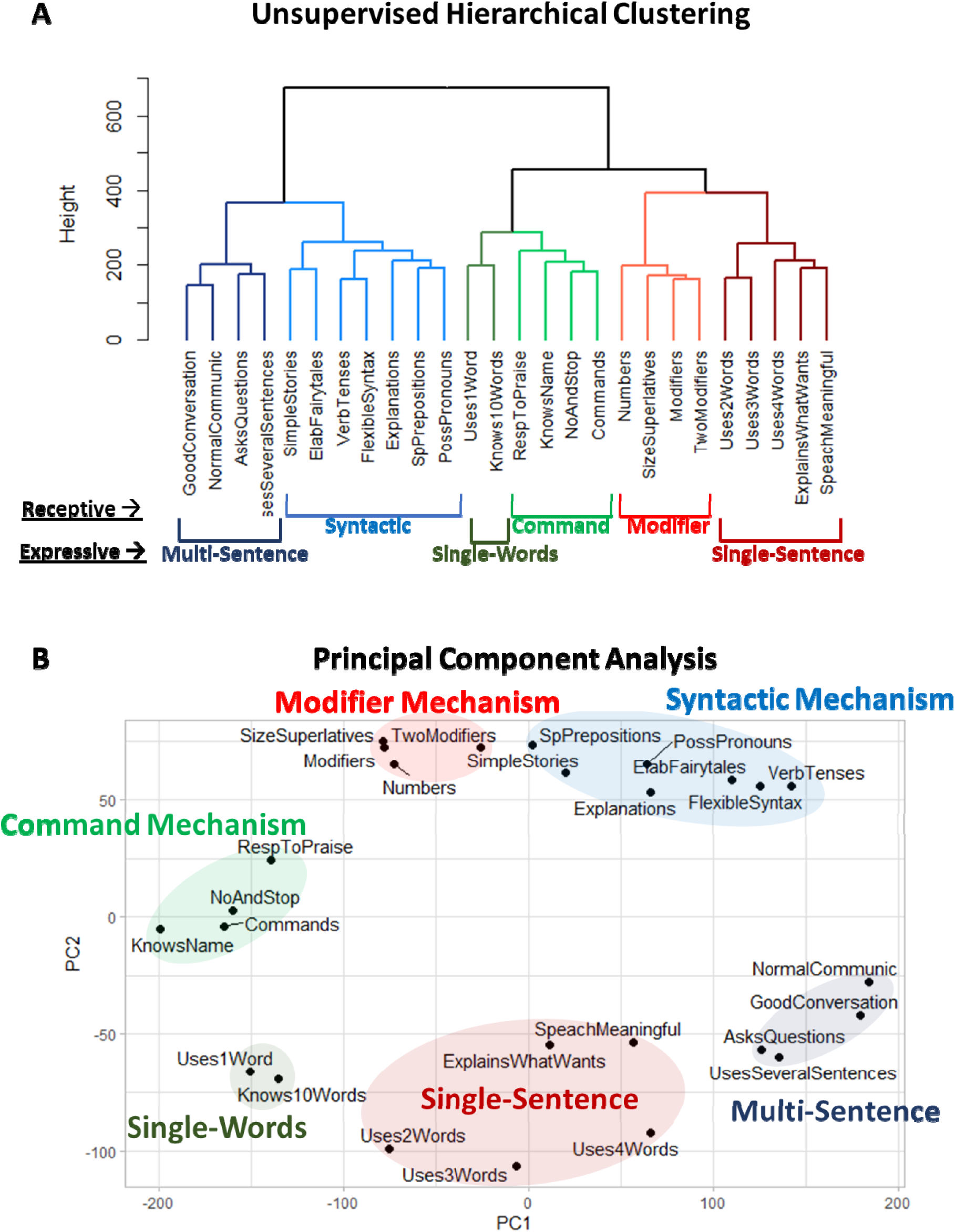
Clustering analysis of 15 receptive language and 11 expressive language items. (A) The dendrogram represents the UHCA of 26 items. (B) PCA of the 26 items shows a clear separation between the six clusters: three language comprehension clusters on the top (Command, Modifier, and Syntactic) and three expressive clusters on the bottom (Single-Word, Single-Sentence, and Multi-Sentence). Principal component 1 accounts for 37.3% of the variance in the data. Principal component 2 accounts for 11.0% of the variance in the data.

As a control, we calculated UHCA and PCA of the 15 language comprehension abilities together with the 11 expressive language items along with the *hyperactivity* item, which is not expected to be related to any particular language ability and therefore should cluster into its own group. As expected, both UHCA and PCA clustered the hyperactivity item into its own group at a significant distance from the six language clusters, validating the effectiveness of both clustering techniques (Figure S3).

### Clustering analysis of receptive-expressive language abilities together with drawing abilities

Next, we calculated UHCA and PCA of the 15 receptive and 11 expressive abilities along with the general drawing item that included non-figurative drawing *“[My child] does drawing, coloring, art*” (Figure 4). Both UHCA and PCA analysis clustered non-figurative drawing with the Modifier Mechanism indicating their common co-occurrence. Both UHCA and PCA clustered the figurative-drawing item *“[My child] draws a VARIETY of RECOGNIZABLE images (objects, people, animals, etc.)*” with the Syntactic Mechanism (Figure 5). Finally, UHCA and PCA grouped the second figurative-drawing item *“[My child] can draw a NOVEL image following YOUR description (e.g. a three-headed horse)*” also with the Syntactic Mechanism (Figure 6).

**Figure 4.**
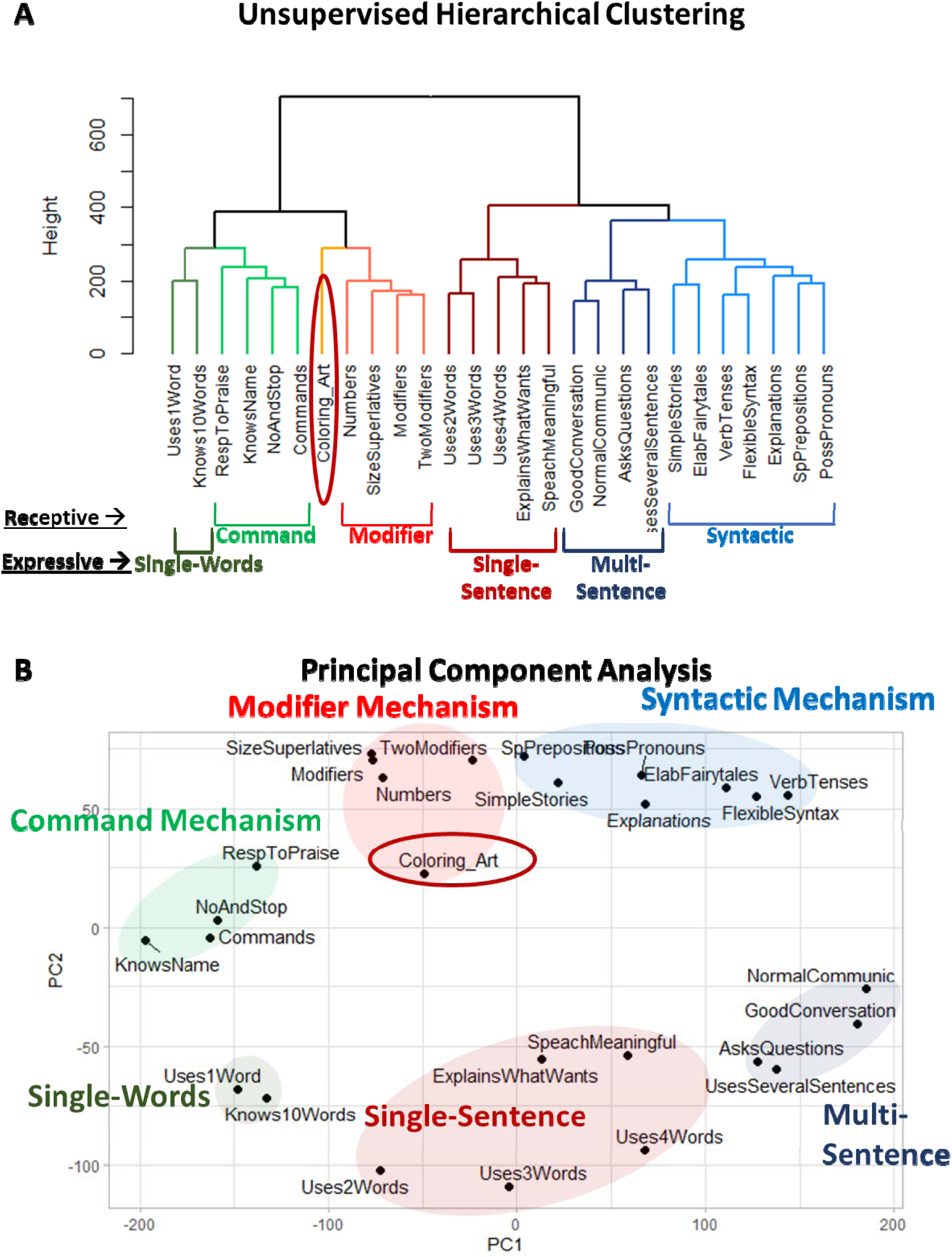
Clustering analysis of 15 receptive language and 11 expressive language items, along with the caregiver-reported item “[My child] does drawing, coloring, art” (labeled *Coloring_Art*). (A) Dendrogram generated using UHCA. (B) PCA, with Principal Component 1 accounting for 36.0% of the variance in the data and Principal Component 2 accounting for 10.6%. The *Coloring_Art* item clusters with the Modifier Mechanism.

**Figure 5.**
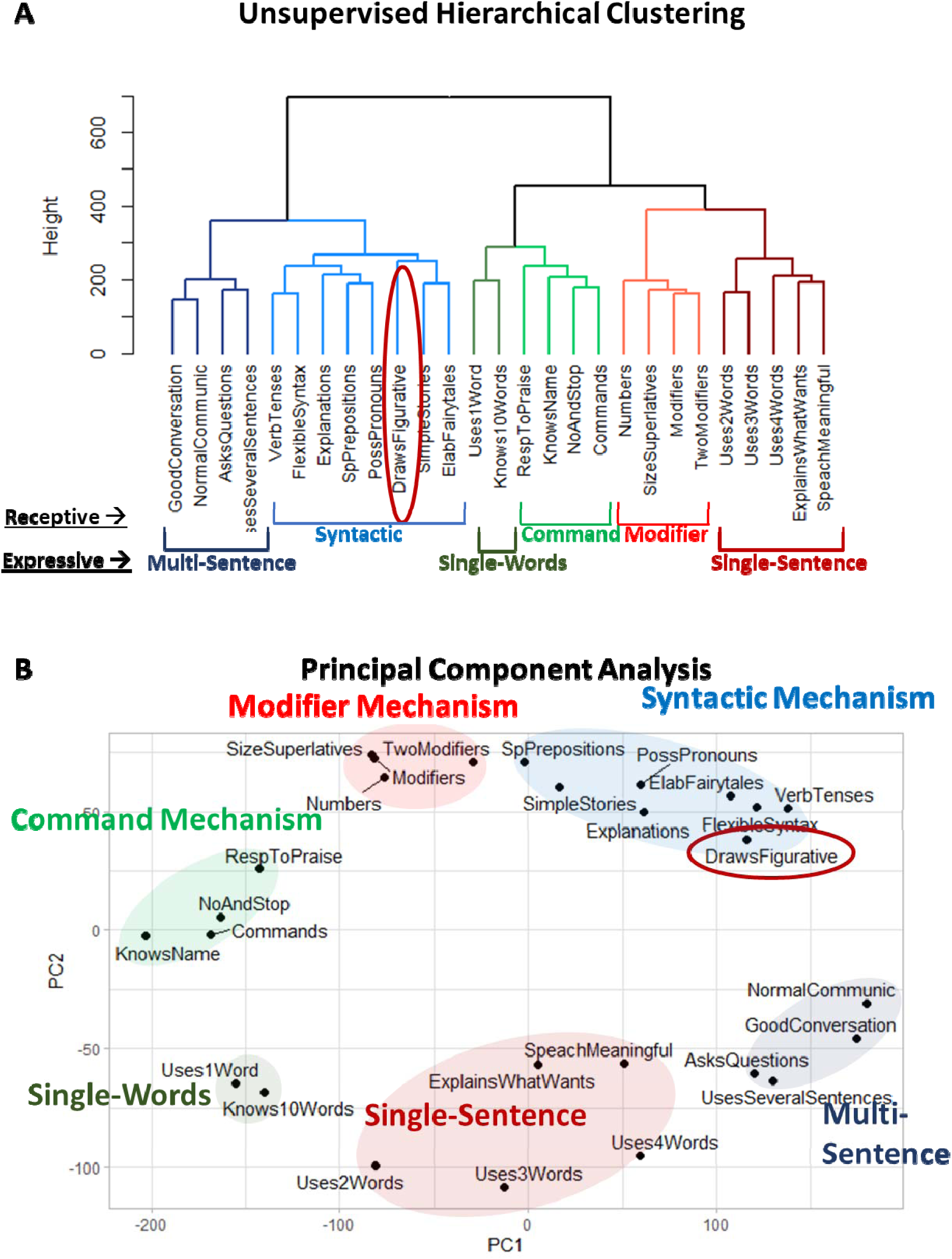
Clustering analysis of 15 receptive language and 11 expressive language items along with the caregiver-reported item “[My child] draws a VARIETY of RECOGNIZABLE images (objects, people, animals, etc.)” labeled *DrawsFigurative*. (A) Dendrogram generated using UHCA. (B) PCA, with Principal Component 1 accounting for 36.9% of the variance in the data and Principal Component 2 accounting for 10.6%. The *DrawsFigurative* item clusters with the Syntactic Mechanism.

**Figure 6.**
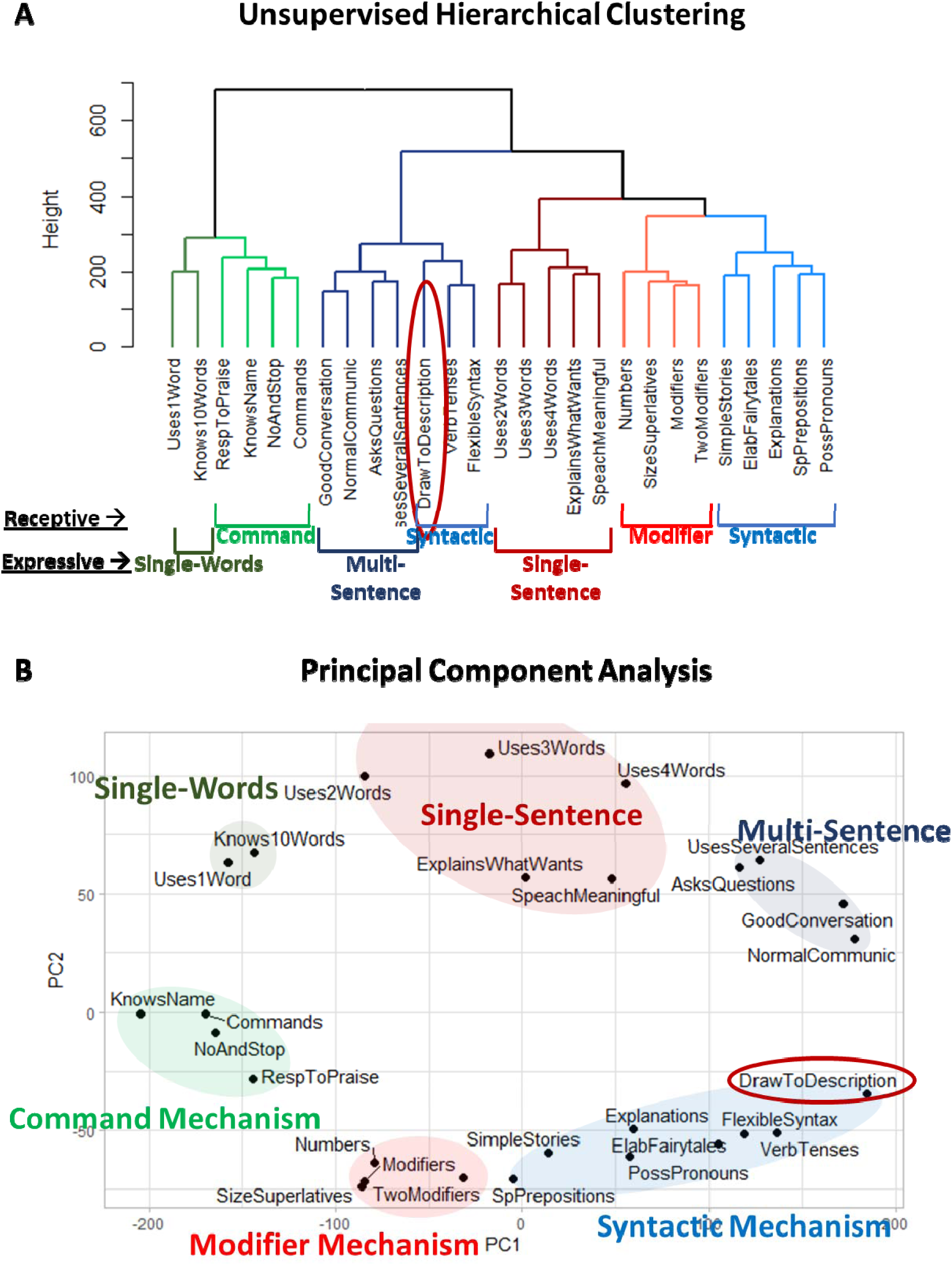
Clustering analysis of 15 receptive language and 11 expressive language items along with the caregiver-reported item “[My child] can draw a NOVEL image following YOUR description (e.g. a three-headed horse)” labeled *DrawToDescription*. (A) Dendrogram generated using UHCA. (B) PCA, with Principal component 1 accounting for 38.5% of the variance in the data and Principal component 2 accounting for 10.5%. The *DrawToDescription* item clusters with the Syntactic Mechanism.

Interpreted in a developmental context, these findings suggest that individuals with language impairments who plateau at the Modifier level typically show a propensity for non-figurative drawing but not for figurative drawing. In contrast, individuals who progress to the Syntactic level usually acquire the ability to draw figuratively.

### Stability of drawing abilities associations with language clusters

We examined the stability of the association between drawing abilities and language clusters across different age groups (4-6 years, n=41,188; 6-12 years, n=32,074; and 12-22 years, n=4,354), levels of autism severity (severe, n=11,498; moderate, n=19,844; and mild, n=31,072), native language (English, n=26,020 and non-English, n=51,596), and participants’ sex (female, n=18,413; male, n=57,398; unknown, n=1,805) (Supplementary Figures 4-31).

Figurative drawing ability, as reported by caregivers in response to the items *“[My child] draws a VARIETY of RECOGNIZABLE images (objects, people, animals, etc.)*” and *“[My child] can draw a NOVEL image following YOUR description (e.g. a three-headed horse)*”, consistently clustered with the Syntactic Mechanism (Supplementary Figures 14-31).

Conversely, the general non-figurative drawing ability, as reported by caregivers in response to the item *“[My child] does drawing, coloring, art*”, never clustered with the Syntactic Mechanism or any of the three expressive mechanisms. PCA consistently positioned this item closer to the Modifier Phenotype (Supplementary Figures 5-13B) and UHCA clustered it with the Modifier Mechanism in most strata (Supplementary Figures 5, 6, 7, 8, 10, 11, and 13A). These results indicate that non-figurative drawing typically emerges in individuals with the Modifier Phenotype. In three subpopulations, UHCA clustered this item with the Command Mechanism (Supplementary Figures 4, 9, 12A), possibly reflecting insufficient resolution of UHCA to detect clustering in smaller subgroups.

## Discussion

The present study asked whether drawing abilities form a single symbolic domain or whether different forms of drawing align with distinct receptive-expressive language abilities. Across 77,616 individuals with neurodevelopmental disorders, caregiver-reported drawing proficiency related more strongly to language comprehension than to expressive language development. Furthermore, non-figurative drawing/coloring/art clustered most closely with the Modifier mechanism, whereas two figurative drawing abilities clustered with the Syntactic mechanism. These associations were observed using two independent multivariate approaches, UHCA and PCA, and were broadly stable across age, sex, autism severity, and native-language subgroups. The findings therefore suggest that drawing abilities are not uniformly related to receptive-expressive language development. Rather, broad non-figurative graphic activity emerges with modifier-level comprehension, whereas figurative drawing is more closely associated with relational-syntactic comprehension.

This pattern is important because figurative drawing requires more than the ability to make intentional marks. To draw a recognizable animal, person, or object, an individual must select diagnostic visual features, organize them into an appropriate spatial configuration, and produce a graphic form that can be interpreted by another observer. Drawing a novel image from verbal description places still stronger demands on the transformation of linguistic input into a structured mental image. These demands plausibly overlap with the relational-combinatorial capacities indexed by the Syntactic mechanism, including the comprehension of spatial relations, event structure, possessive relations, word-order-dependent meanings, stories, and explanations.

When biological abnormalities slow down or prevent full language attainment, receptive and expressive language development follows a distinct step-wise pattern. Receptive comprehension progresses through the Command → Modifier → Syntactic Mechanisms, while expressive language advances through Nonverbal → Single-Word → Single-Sentence → Multi-Sentence production. When development stalls, it typically does so at a full step rather than at an intermediate or half-step ^54^. The results of this study indicate that drawing abilities are strongly associated with specific levels of language comprehension, rather than a stage in expressive language development. The presence of the Syntactic Mechanism is closely linked to figurative drawing: children who do not acquire the Syntactic Mechanism also typically fail to acquire figurative drawing ability. Likewise, the presence of the Modifier Mechanism is strongly associated with non-figurative drawing, and children who do not attain the Modifier Mechanism also usually do not acquire even non-figurative drawing skills.

These results are consistent with data from neurotypical children. Neurotypical children typically acquire the Command Mechanism around age 2, the Modifier Mechanism around age 3, and the Syntactic Mechanism around age 4 years ^56^. Their drawing abilities follow a parallel developmental pattern, with non-figurative scribbling emerging around age 3 and figurative “pre-schematic” drawings appearing by about age 4 ^57^. However, the associations between comprehension and drawing phenotypes are difficult to study in neurotypical populations because children pass through all language levels rapidly and are often introduced to drawing after they acquire the Syntactic Phenotype. In contrast, children with language impairments often remain at inferior levels for years ^54^, providing an ideal population in which to examine the relationship between language mechanisms and drawing abilities.

Relatedly, our non-figurative item combined drawing, coloring, and art into a single caregiver-reported measure, and so it should not be interpreted as a pure index of abstract symbolic mark-making. It may capture interest, exposure, motor control, compliance, caregiver encouragement, or general developmental level, not specifically non-figurative symbolic production. As such, it indexes participation in non-recognizable or non-figurative graphic activity in a broad developmental sense. Future work should aim to refine the dimensions over which to analyze capacities relating to (non-)figurative drawing.

The present findings also have implications for how drawing evidence is interpreted in debates about human cognitive evolution. Archaeological discussions often treat non-figurative marks, hand stencils, ornaments, figurative images, and composite scenes as evidence for a broadly defined symbolic capacity ^58–60^. The present results suggest that this category may be too coarse. In contemporary neurodevelopmental variation, broad non-figurative graphic activity and figurative drawing do not show identical cognitive associations. Non-figurative drawing/coloring/art aligns most closely with Modifier-level comprehension, whereas figurative drawing aligns with relational-Syntactic comprehension.

This does not mean that contemporary neurodevelopmental data can be mapped directly onto the minds of extinct hominins, nor that the archaeological sequence from non-figurative to figurative art can be explained by a single developmental mechanism. The archaeological record is shaped by preservation biases, cultural transmission, ecological context, available materials, demographic structure, and chance discovery. Nevertheless, the dissociation observed here provides a useful constraint on evolutionary interpretation. If figurative depiction depends more strongly than non-figurative mark-making on relational-combinatorial representation, then the later appearance of figurative visual traditions may reflect not only changes in artistic convention or preservation, but also changes in the cognitive operations required to construct, manipulate, and communicate recognizable visual representations.

Under this more cautious interpretation, the transition from non-figurative to figurative art should not be taken as a simple threshold from “non-symbolic” to “symbolic” cognition. Rather, it may reflect a shift among different forms of symbolic and combinatorial capacity. Non-figurative visual production may require intentional mark-making, perceptual salience, and culturally mediated graphic routines. Figurative depiction, especially of novel or composite entities, may additionally require the integration of object knowledge, spatial structure, finely controlled mental imagery, and relational-combinatorial planning. The present findings support this graded view by showing that different drawing abilities associate with different levels of language comprehension in a large neurodevelopmental cohort.

To conclude, syntactic comprehension does not fossilize, making its evolutionary acquisition difficult to determine. Traditionally, it has been examined through the lens of symbolic thinking, which includes drawing ability. This study’s findings suggest that not all drawing abilities depend equally on the syntactic level: figurative drawing is strongly associated with the Syntactic Phenotype, whereas non-figurative drawing is primarily associated with the Modifier Phenotype. This work therefore helps identify a developmental-clinical dissociation in contemporary individuals that may constrain hypotheses about why non-figurative and figurative visual traditions appear at different points in the archaeological record.

## Limitations

The main limitation of this study is its associational nature: a syntactic comprehension phenotype does not directly cause drawing abilities. Some individuals may possess the Syntactic comprehension phenotype but fail to exhibit it in figurative drawing due to limited cultural encouragement or fine-motor constraints. Conversely, others may lack the Syntactic phenotype yet display some figurative drawing ability. Autistic savants are particularly notable in this regard, as their exceptional figurative drawing abilities often contrast sharply with markedly limited syntactic skills ^61,62^. Importantly, the mechanisms supporting figurative drawing in autistic savants may differ from the mental combinatorial pre-planning typically used by neurotypical individuals. For instance, the contours of certain animals can become automatized through extensive practice. In such cases, drawing an animal resembles ‘writing a signature’, executed with minimal conscious planning, enabling drawing of the full outline of the animal in a single fluid motion. In our parent-report assessments, we attempted to mitigate the potential influence of automated drawing by emphasizing ‘*VARIETY of animals’ in the item, “My child draws a VARIETY of RECOGNIZABLE images (objects, people, animals, etc.)*”. However, we cannot determine how many participants in our cohort may have reported the presence of an automatized drawing skill.

Second, the drawing measures were broad caregiver-report items rather than standardized drawing assessments. The general drawing propensity item*—“[My child] does drawing, coloring, art*”—may capture heterogeneous activities, including scribbling, coloring, tracing, craft participation, or exposure to art materials. It should therefore not be interpreted as a pure measure of abstract symbolic mark-making. Similarly, figurative drawing items*—“[My child] draws a VARIETY of RECOGNIZABLE images (objects, people, animals, etc.)*;” and *“[My child] can draw a NOVEL image following YOUR description (e.g. a three-headed horse)*.”—may depend on visual memory, fine-motor control, imitation, practice, cultural exposure, and caregiver expectations, in addition to representational or linguistic abilities.

Third, the Syntactic mechanism label should be interpreted operationally. The items in this cluster include not only word-order-dependent syntax, but also spatial prepositions, verb tense, possessive pronouns, complex explanations, stories, and elaborate fairytales. Thus, the cluster may index a broader capacity for relational-combinatorial comprehension rather than syntax in the narrow grammatical sense. Indeed, the novel-image-from-description item is not a pure drawing measure because it requires comprehension of a verbal prompt. Its association with the Syntactic mechanism may therefore partly reflect the linguistic demands embedded in the item itself.

Fourth, the observed associations may partly reflect general developmental level or item difficulty. Although the differential clustering of non-figurative and figurative drawing items argues against a completely domain-general explanation, additional analyses controlling for age, diagnosis, autism severity, parent education, expressive-language level, and overall developmental severity would further strengthen the interpretation.

Fifth, contemporary neurodevelopmental variation cannot be treated as a direct model of ancestral cognition. The evolutionary implications of the present findings are therefore necessarily indirect. The data show that, in a large contemporary clinical-developmental cohort, non-figurative and figurative drawing abilities have different associations with language-comprehension phenotypes. Any connection to the archaeological sequence of non-figurative and figurative art should be treated as hypothesis-generating rather than definitive.

Finally, some individuals may acquire drawing abilities through atypical routes. For example, highly practiced or savant-like drawing may rely on automatized visuomotor routines rather than the kind of relational-combinatorial planning proposed here. The item requiring a *variety of recognizable images* and the item requiring *a novel image from description* partly mitigate this concern, but they cannot eliminate it.

## Data Availability

De-identified raw data from this manuscript are available from the corresponding author upon reasonable request.

## Acknowledgements

We wish to thank all participants’ caregivers who found time to complete children’s assessments. The language therapy app used to collect the data presented in this manuscript was made possible by the contributions of Rita Dunn, Alexander Faisman, Jonah Elgart, Lisa Lokshina, and Yulia Dumov.

## Author contributions

AV and EK designed the study. RV, EK, and AV analyzed the data. EM and AV wrote the paper.

## References

1. Beyn, E. S. & Knyazeva, G. R. The problem of prosopagnosia. J. Neurol. Neurosurg. Psychiatry 25, 154 (1962).

2. Levine, D. N., Warach, J. & Farah, M. Two visual systems in mental imagery: Dissociation of “what” and “where” in imagery disorders due to bilateral posterior cerebral lesions. Neurology 35, 1010–1010 (1985).

3. Thomas, G. V. & Silk, A. M. An Introduction to the Psychology of Children’s Drawings. (New York University Press, 1990).

4. Zilhão, J. et al. Symbolic use of marine shells and mineral pigments by Iberian Neandertals. Proc. Natl. Acad. Sci. 107, 1023–1028 (2010).

5. Bouzouggar, A. et al. 82,000-year-old shell beads from North Africa and implications for the origins of modern human behavior. Proc. Natl. Acad. Sci. 104, 9964–9969 (2007).

6. Henshilwood, C., d’Errico, F., Vanhaeren, M., Van Niekerk, K. & Jacobs, Z. Middle stone age shell beads from South Africa. Science 304, 404–404 (2004).

7. d’Errico, F., Henshilwood, C., Vanhaeren, M. & Van Niekerk, K. Nassarius kraussianus shell beads from Blombos Cave: evidence for symbolic behaviour in the Middle Stone Age. J. Hum. Evol. 48, 3–24 (2005).

8. Sehasseh, E. M. et al. Early Middle Stone Age personal ornaments from Bizmoune Cave, Essaouira, Morocco. Sci. Adv. 7, eabi8620 (2021).

9. Henshilwood, C. S. et al. An abstract drawing from the 73,000-year-old levels at Blombos Cave, South Africa. Nature 562, 115–118 (2018).

10. Joordens, J. C. et al. Homo erectus at Trinil on Java used shells for tool production and engraving. Nature 518, 228–231 (2015).

11. Marquet, J.-C. et al. The earliest unambiguous neanderthal engravings on cave walls: La Roche-Cotard, Loire valley, France. PLoS One 18, e0286568 (2023).

12. Henshilwood, C. S., d’Errico, F. & Watts, I. Engraved ochres from the middle stone age levels at Blombos Cave, South Africa. J. Hum. Evol. 57, 27–47 (2009).

13. Hoffmann, D. L. et al. U-Th dating of carbonate crusts reveals Neandertal origin of Iberian cave art. Science 359, 912–915 (2018).

14. Standish, C. D. et al. The age of hand stencils in Maltravieso cave (Extremadura, Spain) established by U-Th dating, and its implications for the early development of art. J. Archaeol. Sci. Rep. 61, 104891 (2025).

15. Pettitt, P. et al. Are hand stencils in European cave art older than we think? An evaluation of the existing data and their potential implications. Prehist. Art Prehist. Cult. Stud. Honour Profr. Rodrigo Balbín-Behrmann 31–43 (2015).

16. Navarro, V. F., Casares, D. F., Martínez, D. G. & Maidagan, D. G. Decoding Palaeolithic Hand Stencils: Age and Sex Identification Through Geometric Morphometrics. J. Archaeol. Method Theory 32, 24 (2025).

17. Ripoll, S. et al. Hands Stencils in El Castillo Cave (Puente Viesgo, Cantabria, Spain). An Interdisciplinary Study. in Proceedings of the prehistoric society vol. 87 51–71 (Cambridge University Press, 2021).

18. Fernández-Navarro, V., Spaey, O. & Garate, D. Reevaluating Hand Stencil Phenomena in Cave Art: A Step Forward towards the Characterization of Symbolic Patterns during the Upper Palaeolithic in Europe. Camb. Archaeol. J. 1, 21 (2026).

19. Aubert, M. et al. Earliest hunting scene in prehistoric art. Nature 576, 442–445 (2019).

20. Aubert, M. et al. Palaeolithic cave art in Borneo. Nature 564, 254–257 (2018).

21. Aubert, M. et al. Pleistocene cave art from Sulawesi, Indonesia. Nature 514, 223–227 (2014).

22. Bourrillon, R., et al. A new Aurignacian engraving from Abri Blanchard, France: Implications for understanding Aurignacian graphic expression in Western and Central Europe. Quat. Int. http://www.sciencedirect.com/science/article/pii/S1040618216305766 (2017).

23. Dalton, R. Lion Man Takes Pride of Place as Oldest Statue. (Nature Publishing Group, 2003).

24. Moreno Campos, V. & Benítez-Burraco, A. Communication deficits in a case of a deletion in 7q31.1-q31.33 encompassing FOXP2. Clin. Linguist. Phon. 37, 1157–1170 (2023).

25. Morison, L. D., Braden, R., Amor, D. J. & Morgan, A. T. Speech and Language Disorders Associated With 7q31 Deletions Implicating FOXP2. Am. J. Med. Genet. A. e64190 (2025) doi:10.1002/ajmg.a.64190.

26. Kuo, H.-Y. et al. Speech-and language-linked FOXP2 mutation targets protein motors in striatal neurons. Brain 146, 3542–3557 (2023).

27. Martins, P. T., Marí, M. & Boeckx, C. SRGAP2 and the gradual evolution of the modern human language faculty. J. Lang. Evol. 3, 67–78 (2018).

28. Toma, C. et al. Analysis of two language-related genes in autism: A case–control association study of: FOXP2: And: CNTNAP2. Psychiatr. Genet. 23, 82–85 (2013).

29. Vyshedskiy, A., Venkatesh, R., Khokhlovich, E. & Satik, D. Three mechanisms of language comprehension are revealed through cluster analysis of individuals with language deficits. Npj Sci. Learn. 9, 1–12 (2024).

30. Vyshedskiy, A., Venkatesh, R. & Khokhlovich, E. Are there distinct levels of language comprehension in autistic individuals – cluster analysis. Npj Ment. Health Res. 3, (2024).

31. Murphy, E., Venkatesh, R., Khokhlovich, E. & Vyshedskiy, A. Merge-based syntax is mediated by distinct neurocognitive mechanisms in 84,000 individuals with language deficits across nine languages. Sci. Rep. 15, 44444 (2025).

32. Gankin, K., Venkatesh, R., Khokhlovich, E. & Vyshedskiy, A. Receptive and expressive language phenotyping in over 62,000 autistic Individuals. https://www.researchsquare.com/article/rs-8466583/latest (2026).

33. Vyshedskiy, A. & Dunn, R. Mental Imagery Therapy for Autism (MITA)-An Early Intervention Computerized Brain Training Program for Children with ASD. Autism Open Access 5, 2 (2015).

34. Dunn, R. et al. Comparison of performance on verbal and nonverbal multiple-cue responding tasks in children with ASD. Autism Open Access 7, 218 (2017).

35. Dunn, R. et al. Tablet-Based Cognitive Exercises as an Early Parent-Administered Intervention Tool for Toddlers with Autism - Evidence from a Field Study. Clin. Psychiatry 3, (2017).

36. Dunn, R. et al. Children With Autism Appear To Benefit From Parent-Administered Computerized Cognitive And Language Exercises Independent Of the Child’s Age Or Autism Severity. Autism Open Access 7, (2017).

37. Vyshedskiy, A. et al. Novel prefrontal synthesis intervention improves language in children with autism. Healthcare 8, 566 (2020).

38. Rimland, B. & Edelson, S. M. Autism treatment evaluation checklist (ATEC). Autism Res. Inst. San Diego CA http://www.autism.com (1999).

39. Braverman, J., Dunn, R. & Vyshedskiy, A. Development of the Mental Synthesis Evaluation Checklist (MSEC): A Parent-Report Tool for Mental Synthesis Ability Assessment in Children with Language Delay. Children 5, 62 (2018).

40. Fridberg, E., Khokhlovich, E. & Vyshedskiy, A. Watching Videos and Television Is Related to a Lower Development of Complex Language Comprehension in Young Children with Autism. in Healthcare vol. 9 423 (Multidisciplinary Digital Publishing Institute, 2021).

41. Acosta, A., Khokhlovich, E., Reis, H. & Vyshedskiy, A. Dietary factors impact developmental trajectories in young autistic children. J. Autism Dev. Disord. 54, 3533–3548 (2023).

42. Forman, P., Khokhlovich, E. & Vyshedskiy, A. Longitudinal Developmental Trajectories in Young Autistic Children Presenting with Seizures, Compared to those Presenting without Seizures, Gathered via Parent-report Using a Mobile Application. J. Dev. Phys. Disabil. 10.1007/s10882-022-09851-y (2022) doi:10.1007/s10882-022-09851-y.

43. Benítez-Burraco, A., Hoshi, K. & Murphy, E. Language deficits in GRIN2A mutations and Landau–Kleffner syndrome as neural dysrhythmias. J. Neurolinguistics 67, 101139 (2023).

44. Levin, J., Khokhlovich, E. & Vyshedskiy, A. Longitudinal developmental trajectories in young autistic children presenting with sleep problems, compared to those presenting without sleep problems, gathered via parent-report using a mobile application. Res. Autism Spectr. Disord. 97, 102024 (2022).

45. Arnold, M. & Vyshedskiy, A. Combinatorial language parent-report score differs significantly between typically developing children and those with Autism Spectrum Disorders. J. Autism Dev. Disord. 10.1007/s10803-022-05769-8 (2022) doi:/10.1007/s10803-022-05769-8.

46. American Psychiatric Association. Diagnostic and Statistical Manual of Mental Disorders (DSM-5®). (American Psychiatric Pub, 2013).

47. Jagadeesan, P., Kabbani, A. & Vyshedskiy, A. Parent-reported assessment scores reflect ASD severity level in 2- to 7- year-old children. Children 9, 701 (2022).

48. World Medical Association. World Medical Association Declaration of Helsinki: ethical principles for medical research involving human subjects. JAMA 310, 2191–2194 (2013).

49. R Foundation for Statistical Computing. R: A language and environment for statistical computing. (2021).

50. Robitzsch, A. Why ordinal variables can (almost) always be treated as continuous variables: Clarifying assumptions of robust continuous and ordinal factor analysis estimation methods. in Frontiers in education vol. 5 589965 (Frontiers Media SA, 2020).

51. Norman, G. Likert scales, levels of measurement and the “laws” of statistics. Adv. Health Sci. Educ. 15, 625–632 (2010).

52. Muthén, B. & Kaplan, D. A comparison of some methodologies for the factor analysis of non-normal Likert variables. Br. J. Math. Stat. Psychol. 38, 171–189 (1985).

53. Rhemtulla, M., Brosseau-Liard, P. É. & Savalei, V. When can categorical variables be treated as continuous? A comparison of robust continuous and categorical SEM estimation methods under suboptimal conditions. Psychol. Methods 17, 354 (2012).

54. Venkatesh, R., Nowakowski, A., Khokhlovich, E. & Vyshedskiy, A. Longitudinal trajectories across the Command, Modifier, and Syntactic Phenotypes of language comprehension in over 6,000 autistic children. Preprint at 10.64898/2025.12.19.25342690 (2025).

55. Venkatesh, R., Khokhlovich, E. & Vyshedskiy, A. Transition dynamics between the four receptive-expressive language phenotypes in autism: Nonverbal, Single-Word, Single-Sentence, and Multi-Sentence production (manuscript in preparation). (2026).

56. Vyshedskiy, A. et al. Language Comprehension Developmental Milestones in Typically Developing Children Assessed by the New Language Phenotype Assessment (LPA). Child. Basel Switz. 12, 793 (2025).

57. Piaget, J. Play, Dreams and Imitation in Childhood. (W. W. Norton & Co. Orig.: La formation du symbole chez l’enfant. Neuchatel: Delachaux & Niestlé., 1945).

58. Tylén, K. et al. The evolution of early symbolic behavior in Homo sapiens. Proc. Natl. Acad. Sci. 117, 4578–4584 (2020).

59. Wisher, I. et al. Beyond the image: Interdisciplinary and contextual approaches to understanding symbolic cognition in Paleolithic parietal art. Evol. Anthropol. 32, 256–259 (2023).

60. Murphy, E. No country for Oldowan men: emerging factors in language evolution. Front. Psychol. 10, 1448 (2019).

61. Pring, L., Ryder, N., Crane, L. & Hermelin, B. Creativity in savant artists with autism. Autism 16, 45–57 (2012).

62. Sacks, O. The Man Who Mistook His Wife for a Hat: A Revealing Exploration of the Mysteries of the Human Mind. vol. 19 (Pan Macmillan, 2014).

