## Supplemental material for "Figurative Drawing Ability is Associated with Advanced Compositional Language Comprehension in Neurodevelopmental Disorders"

### Supplementary Material

Supplementary Table 1: Autism Treatment Evaluation Checklist (ATEC) ^1^, subscale 1: Speech/Language/Communication. The answers choices were: not true, somewhat true, very true.

**1. Knows own name**

**2. Responds to ‘No’ or ‘Stop’**

**3. Can follow some commands**

4. Can use one word at a time (No!, Eat, Water, etc.)

5. Can use 2 words at a time (Don't want, Go home)

6. Can use 3 words at a time (Want more milk)

7. Knows 10 or more words

8. Can use sentences with 4 or more words

9. Explains what he/she wants

10. Asks meaningful questions

11. Speech tends to be meaningful/relevant

12. Often uses several successive sentences

13. Carries on fairly good conversation

14. Has normal ability to communicate for his/her age

Supplementary Table 2: ATEC subscale 2: Sociability**.** The answers choices were: not true, somewhat true, very true.

1. Seems to be in a shell – you cannot reach him/her

2. Ignores other people

3. Pays little or no attention when addressed

4. Uncooperative and resistant

5. No eye contact

6. Prefers to be left alone

7. Shows no affection

8. Fails to greet parents

9. Avoids contact with others

10. Does not imitate

11. Dislikes being held/cuddled

12. Does not share or show

13. Does not wave ‘bye bye’

14. Disagreeable/not compliant

15. Temper tantrums

16. Lacks friends/companions

17. Rarely smiles

18. Insensitive to other's feelings

19. Indifferent to being liked

20. Indifferent if parent(s) leave

Supplementary Table 3: ATEC subscale 3: Sensory/Cognitive awareness**.** The answers choices were: not true, somewhat true, very true.

1. Responds to own name

**2. Responds to praise**

3. Looks at people and animals

4. Looks at pictures (and T.V.)

5. Does drawing, coloring, art

6. Plays with toys appropriately

7. Appropriate facial expression

8. Understands stories on T.V.

9. Understands explanations

10. Aware of environment

11. Aware of danger

12. Shows imagination

13. Initiates activities

14. Dresses self

15. Curious, interested

16. Venturesome - explores

17. “Tuned in” — Not spacey

18. Looks where others are looking

Supplementary Table 4: ATEC subscale 4: Health/Physical/Behavior**.** The answers choices were: not a problem, minor problem, moderate problem, and serious problem.

1. Bed-wetting

2. Wets pants/diapers

3. Soils pants/diapers

4. Diarrhea

5. Constipation

6. Sleep problems

7. Eats too much/too little

8. Extremely limited diet

9. Hyperactive

10. Lethargic

11. Hits or injures self

12. Hits or injures others

13. Destructive

14. Sound-sensitive

15. Anxious/fearful

16. Unhappy/crying

17. Seizures

18. Obsessive speech

19. Rigid routines

20. Shouts or screams

21. Demands sameness

22. Often agitated

23. Not sensitive to pain

24. “Hooked” or fixated on certain objects/topics

25. Repetitive movements (stimming, rocking, etc.)

Supplementary Table 5: Mental Synthesis Evaluation Checklist (MSEC) ^2^. The answers choices were: not true, somewhat true, very true.

**1. Understands simple stories that are read aloud**

**2. Understands elaborate fairy tales that are read aloud (i.e. stories describing FANTASY creatures)**

3. Draws a VARIETY of RECOGNIZABLE images (objects, people, animals, etc.)

4. Can draw a NOVEL image following YOUR description (e.g. a three-headed horse)

5. Engages in a VARIETY of make-believe activities (such as: playing house, playing with toy soldiers, building forts and castles, etc.)

**6. Understands some simple modifiers (i.e. green apple vs. red apple or big apple vs. small apple)**

**7. Understands several modifiers in a sentence (i.e. small green apple)**

**8. Understands size (can select the largest/smallest object out of a collection of objects)**

**9. Understands possessive pronouns (i.e. your apple vs. her apple)**

**10. Understands spatial prepositions (i.e. put the apple ON TOP of the box vs. INSIDE the box vs. BEHIND the box)**

**11. Understands verb tenses (i.e. I will eat an apple vs. I ate an apple)**

**12. Understands the change in meaning when the order of words is changed (i.e. understands the difference between 'a cat ate a mouse' vs. 'a mouse ate a cat')**

**13. Understands NUMBERS (i.e. two apples vs. three apples)**

14. Can perform simple arithmetic: 2 + 3 = ?

15. Can add larger numbers: 7 + 6 = ?

16. Can perform simple subtraction: 3 – 2 = ?

17. Can subtract larger numbers: 15 – 7 = ?

18. Can perform simple multiplication: 2 × 2 = ?

19. Can multiply larger numbers: 6 × 7 = ?

**20. Understands explanations about people, objects or situations beyond the immediate surroundings (e.g., “Mom is walking the dog,” “The snow has turned to water”)**

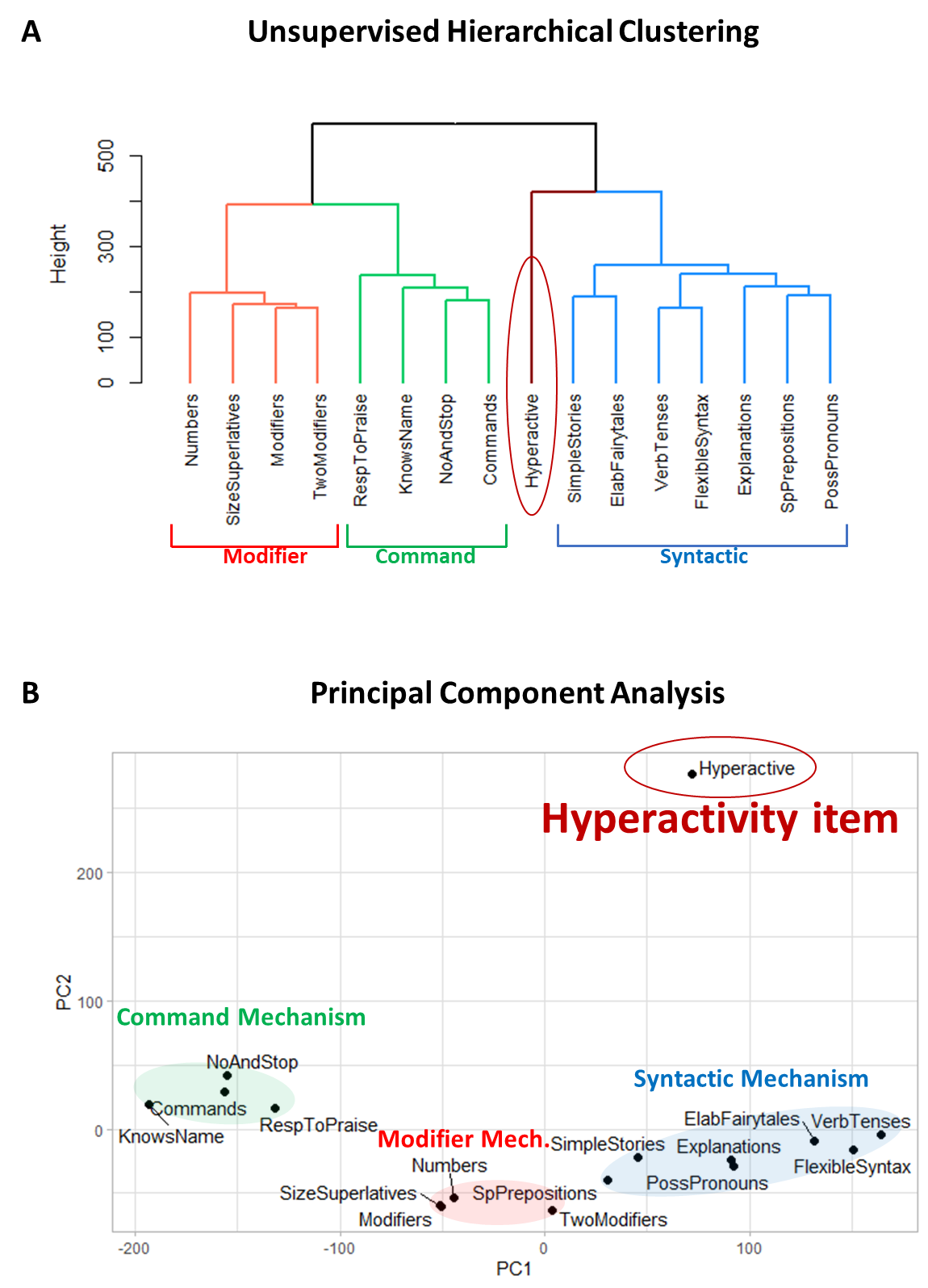

Supplementary Figure 1. Clustering analysis of 15 language comprehension items with the Hyperactivity item. (A) The dendrogram representing the unsupervised hierarchical clustering of language comprehension abilities. (B) Principal component analysis. Principal component 1 accounts for 35.4% of the variance in the data. Principal component 2 accounts for 17.1% of the variance in the data.

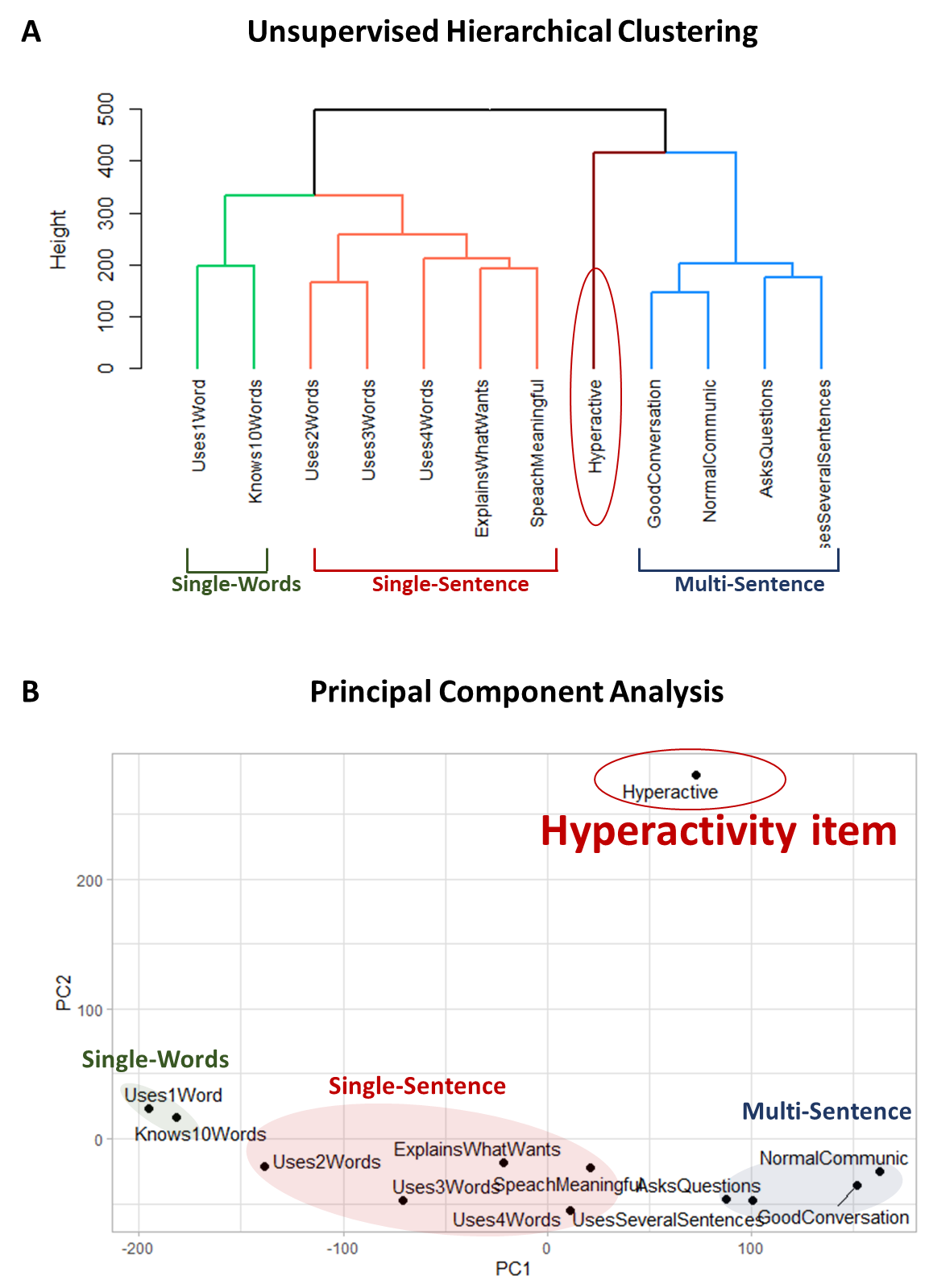

Supplementary Figure 2. The clustering solution of the 11 expressive language abilities along with the “hyperactivity” item. (A) The dendrogram representing the hierarchical clustering. (B) Principal component analysis. Principal component 1 accounts for 40.5% of the variance in the data. Principal component 2 accounts for 22.1% of the variance in the data. The hyperactivity item clusters into its own group at a significant distance from the three language clusters, confirming that hyperactivity item is not related to expressive language abilities.

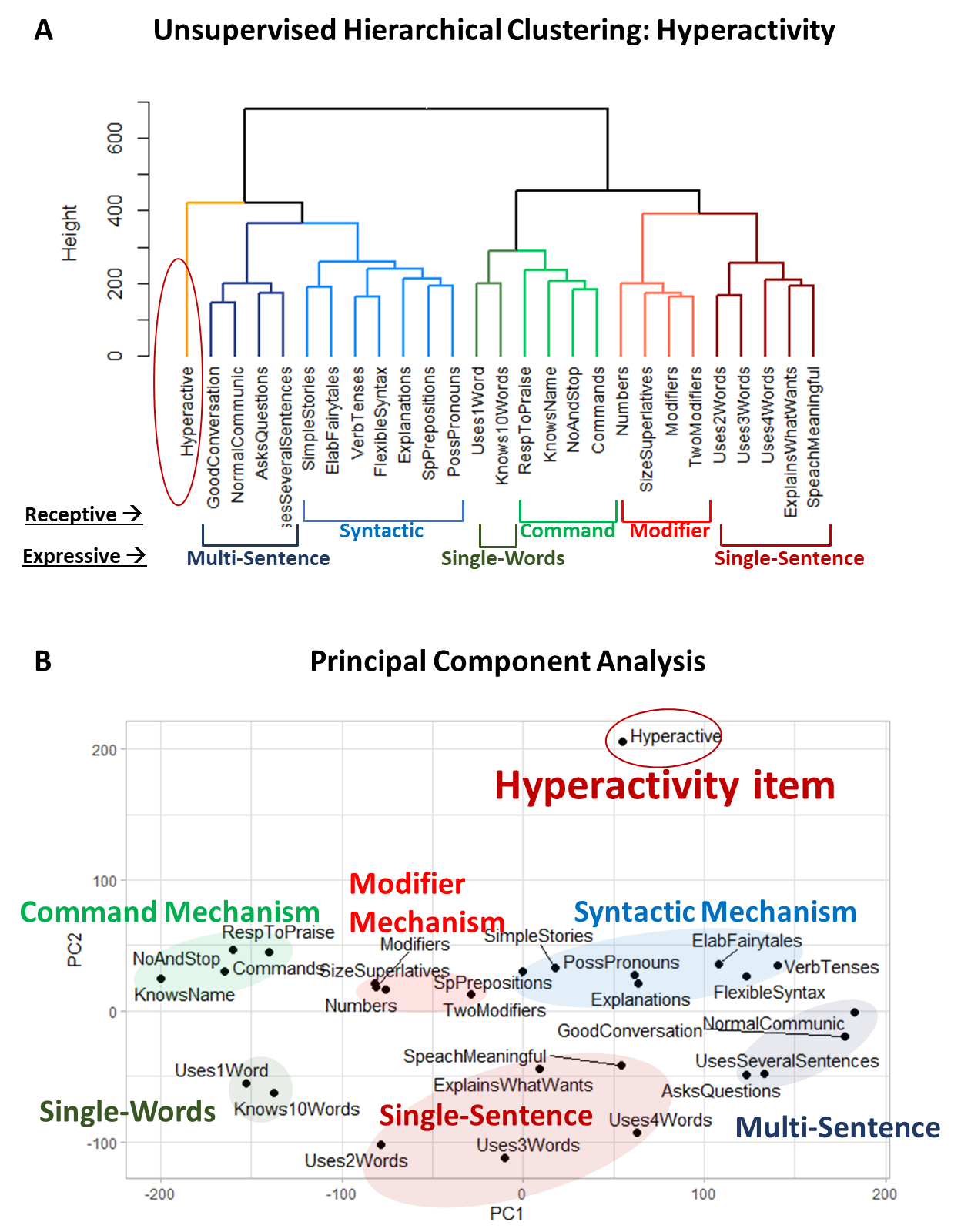
Supplementary Figure 3. The clustering solution of the 15 comprehension and 11 expressive abilities along with the “hyperactivity” item. (A) The dendrogram representing the hierarchical clustering. (B) Principal component analysis. Principal component 1 accounts for 34.1% of the variance in the data. Principal component 2 accounts for 10.2% of the variance in the data. The hyperactivity item clusters into its own group at a significant distance from the three language clusters, confirming that hyperactivity item is not related to language abilities.

### Clustering analysis of the caregiver-reported item “[My child] does drawing, coloring, art”

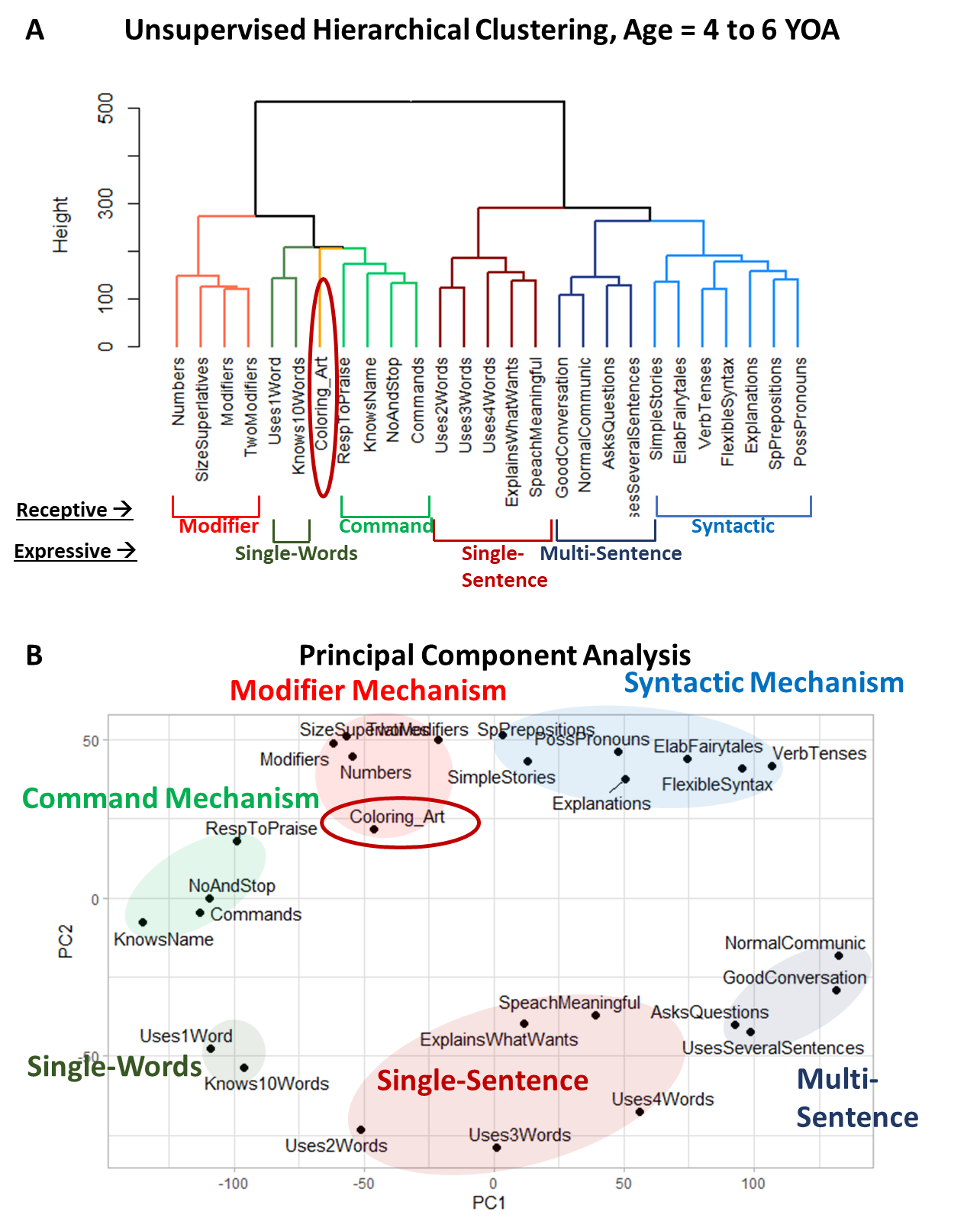
Supplementary Figure 4. Clustering analysis of 15 receptive language and 11 expressive language items, along with the caregiver-reported item “[My child] does drawing, coloring, art” (labeled *Coloring_Art*) in participants aged 4 to 6 years. (A) Dendrogram generated using UHCA. (B) PCA, with Principal Component 1 accounting for 35.6% of the variance in the data and Principal Component 2 accounting for 10.5%.

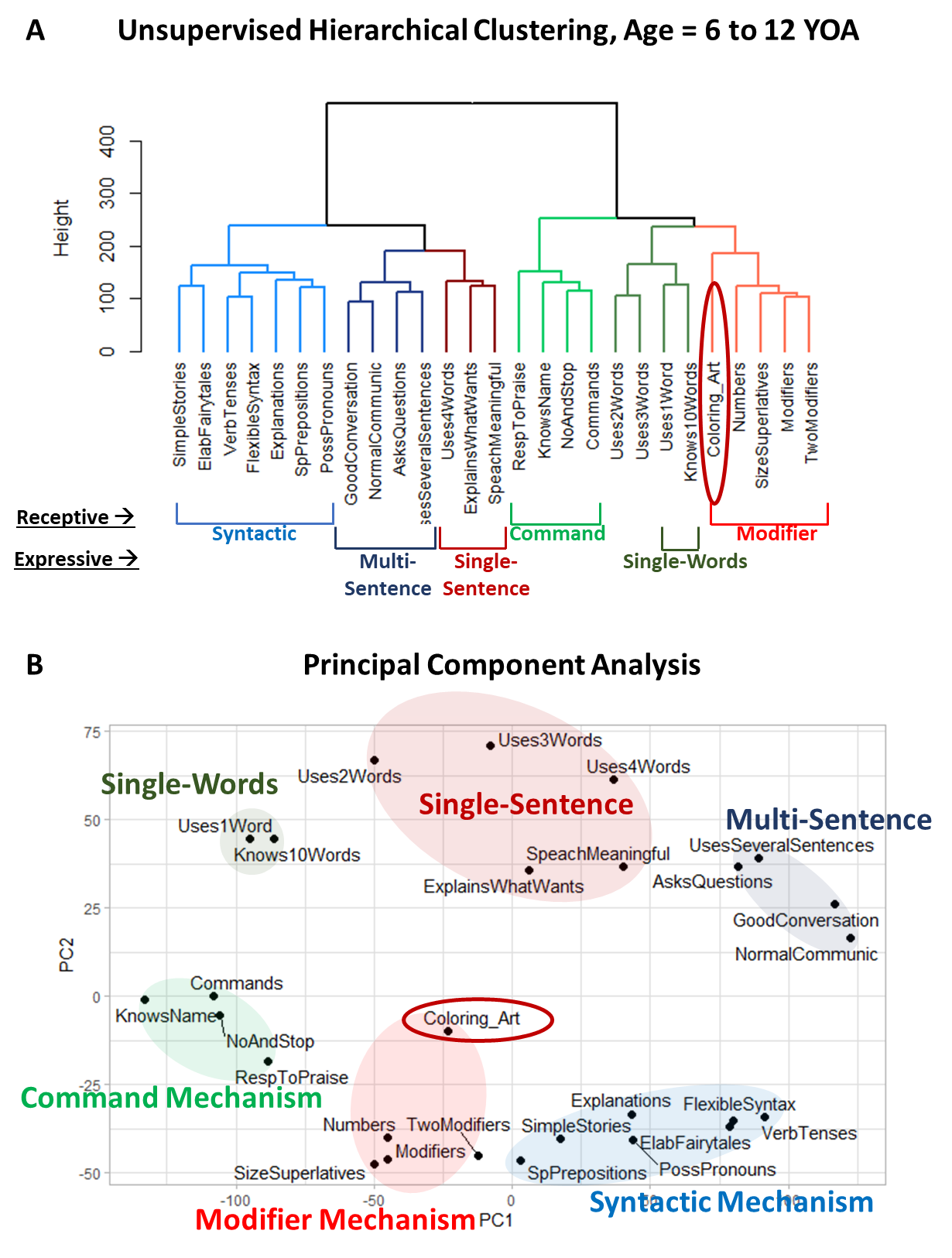
Supplementary Figure 5. Clustering analysis of 15 receptive language and 11 expressive language items, along with the caregiver-reported item “[My child] does drawing, coloring, art” (labeled *Coloring_Art*) in participants aged 6 to 12 years. (A) Dendrogram generated using UHCA. (B) PCA, with Principal Component 1 accounting for 36.8% of the variance in the data and Principal Component 2 accounting for 10.7%.

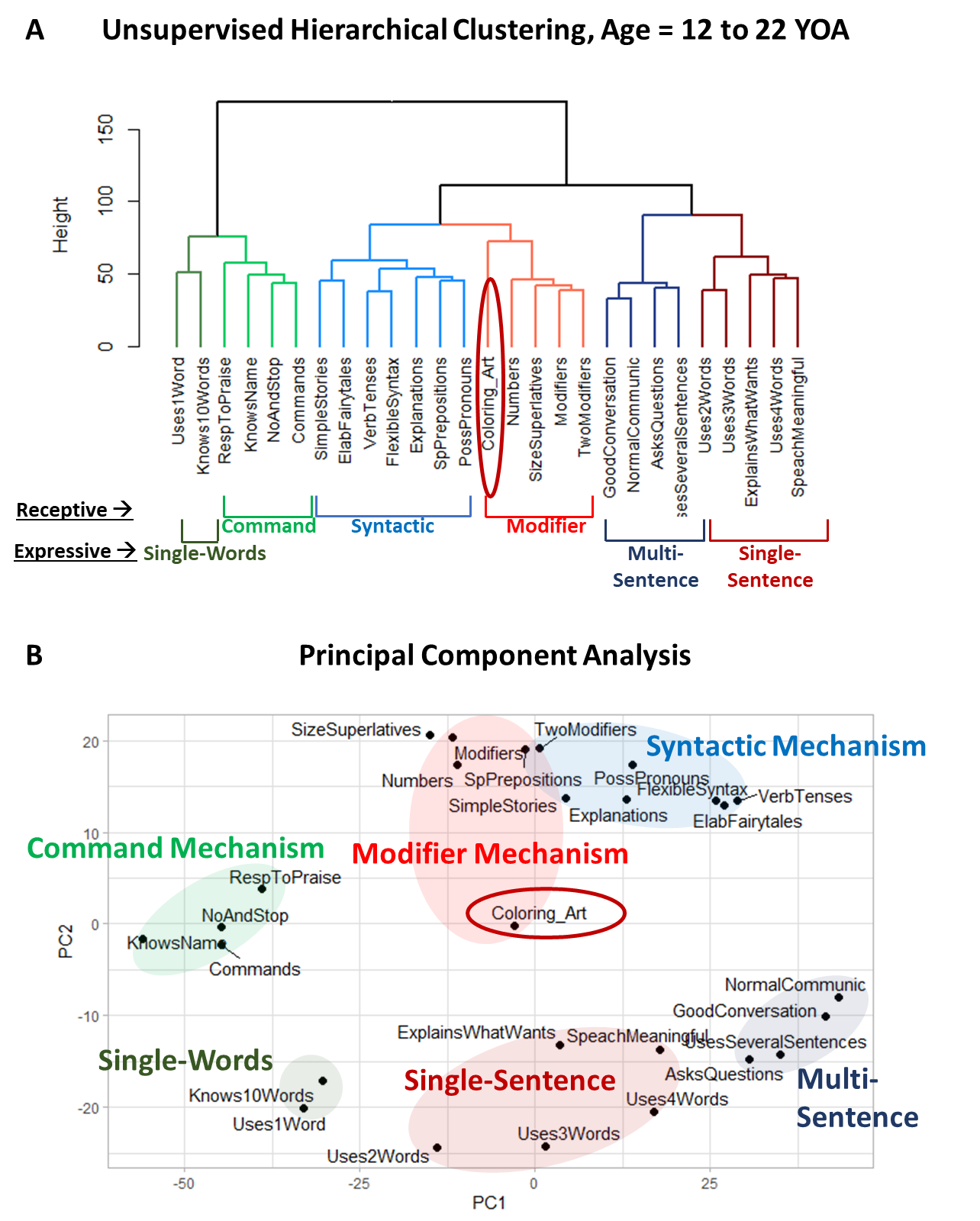
Supplementary Figure 6. Clustering analysis of 15 receptive language and 11 expressive language items, along with the caregiver-reported item “[My child] does drawing, coloring, art” (labeled *Coloring_Art*) in participants aged 12 to 22 years. (A) Dendrogram generated using UHCA. (B) PCA, with Principal Component 1 accounting for 36.6% of the variance in the data and Principal Component 2 accounting for 11.4%.

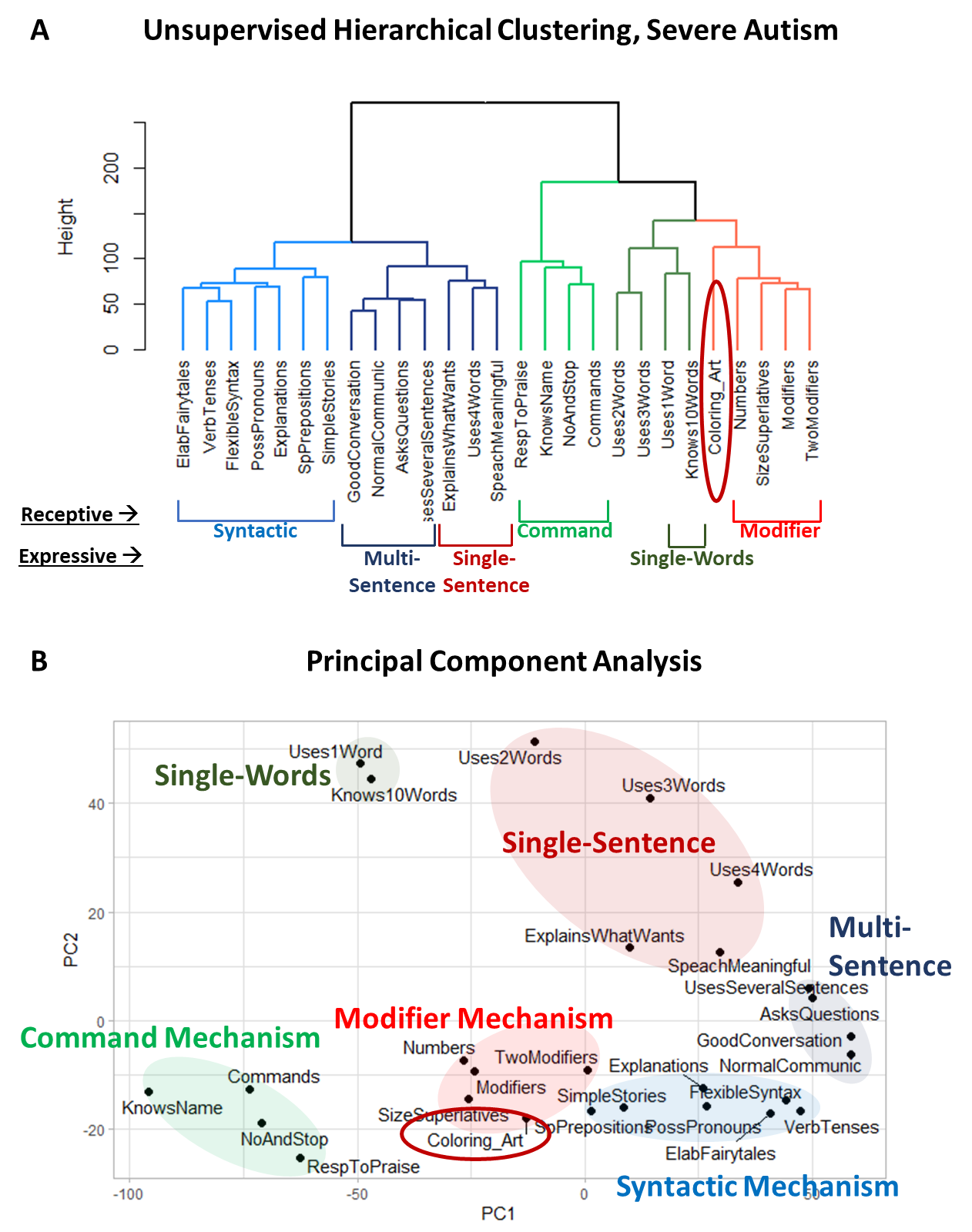

Supplementary Figure 7. Clustering analysis of 15 receptive language and 11 expressive language items, along with the caregiver-reported item “[My child] does drawing, coloring, art” (labeled *Coloring_Art*) in participants diagnosed with severe autism. (A) Dendrogram generated using UHCA. (B) PCA, with Principal Component 1 accounting for 38% of the variance in the data and Principal Component 2 accounting for 9.8%.

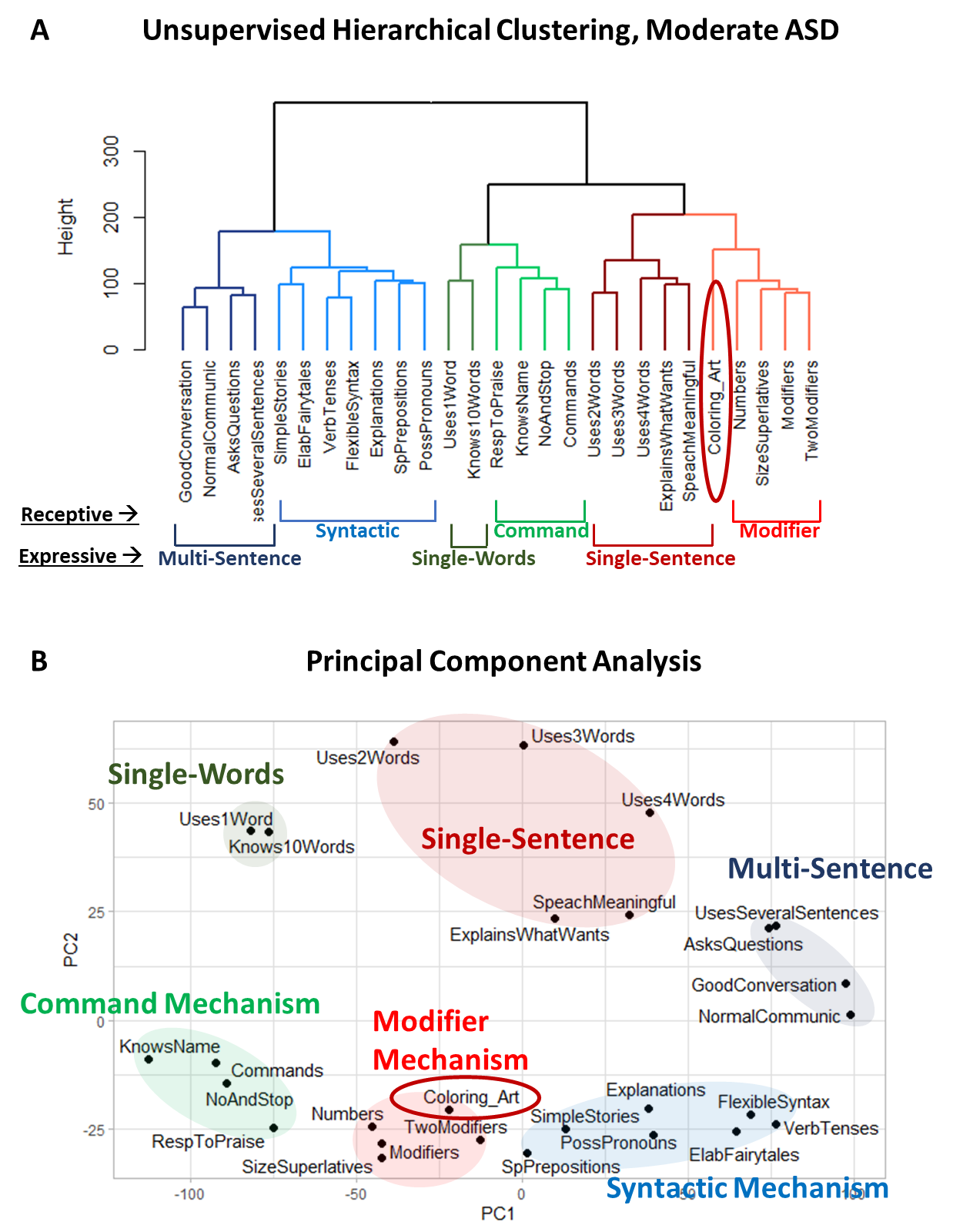
Supplementary Figure 8. Clustering analysis of 15 receptive language and 11 expressive language items, along with the caregiver-reported item “[My child] does drawing, coloring, art” (labeled *Coloring_Art*) in participants diagnosed with moderate autism. (A) Dendrogram generated using UHCA. (B) PCA, with Principal Component 1 accounting for 40.5% of the variance in the data and Principal Component 2 accounting for 9.6%.

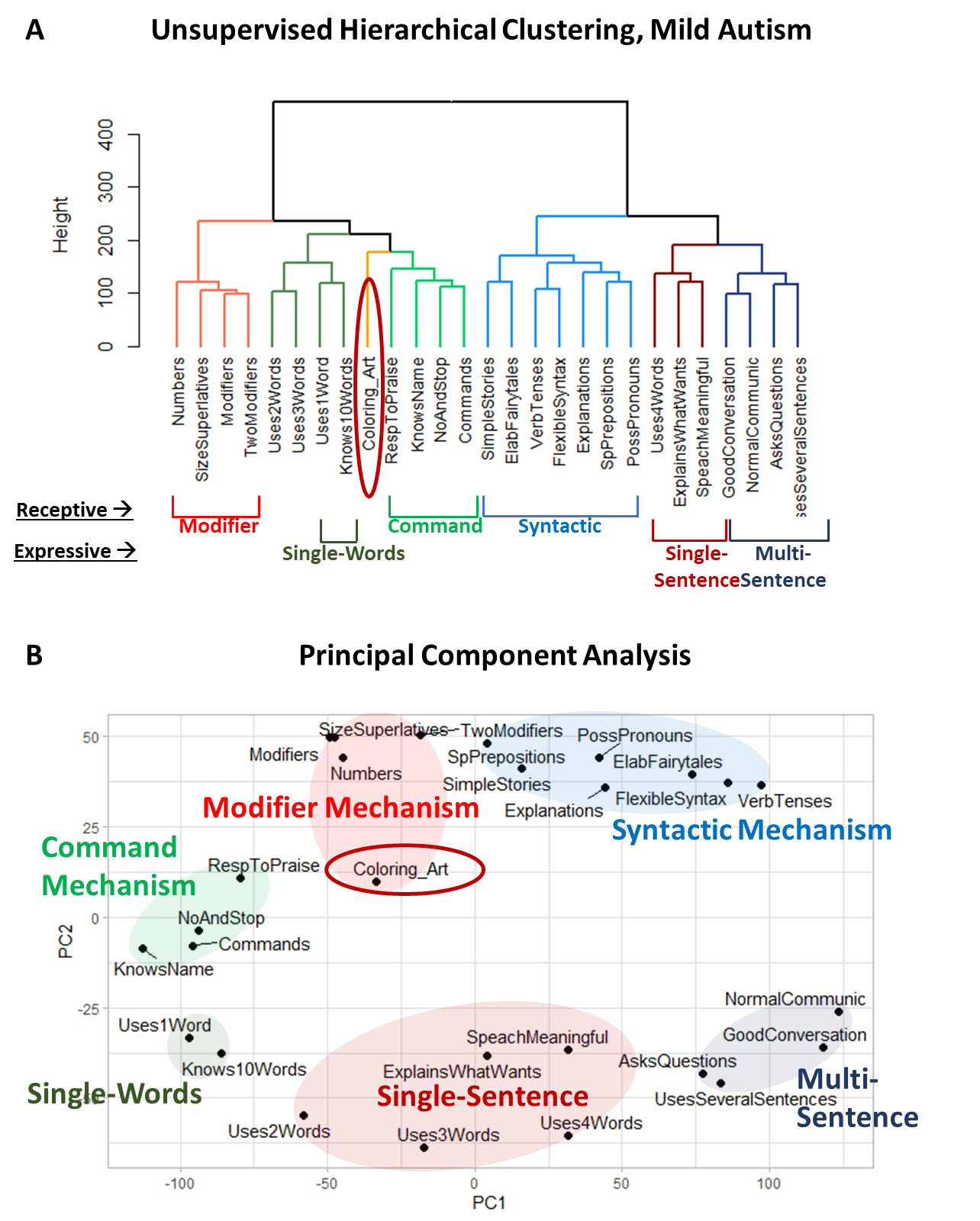
Supplementary Figure 9. Clustering analysis of 15 receptive language and 11 expressive language items, along with the caregiver-reported item “[My child] does drawing, coloring, art” (labeled *Coloring_Art*) in participants diagnosed with mild autism. (A) Dendrogram generated using UHCA. (B) PCA, with Principal Component 1 accounting for 35.6% of the variance in the data and Principal Component 2 accounting for 11.3%.

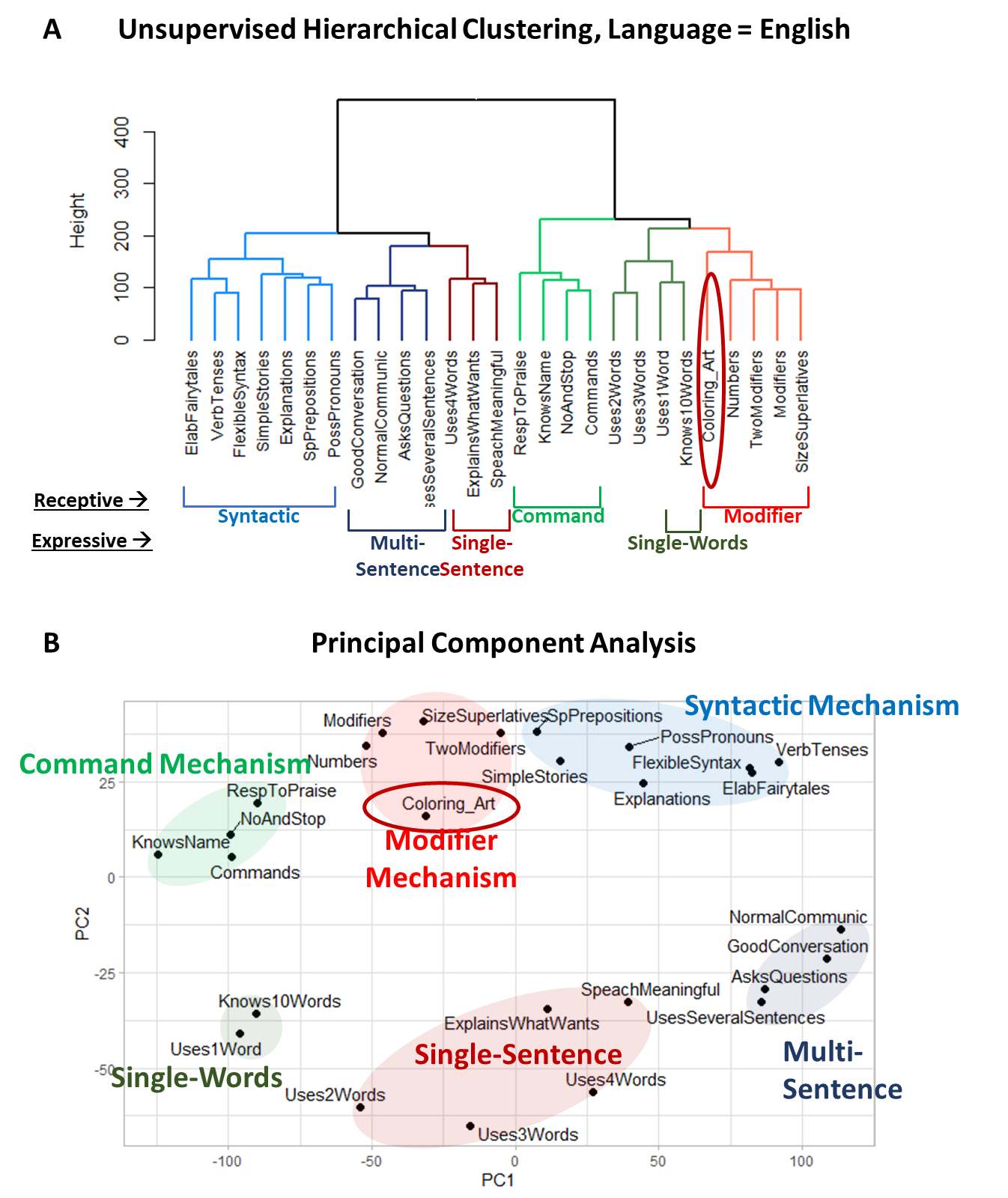
Supplementary Figure 10. Clustering analysis of 15 receptive language and 11 expressive language items, along with the caregiver-reported item “[My child] does drawing, coloring, art” (labeled *Coloring_Art*) in English-speaking participants. (A) Dendrogram generated using UHCA. (B) PCA, with Principal Component 1 accounting for 41.7% of the variance in the data and Principal Component 2 accounting for 9.6%.

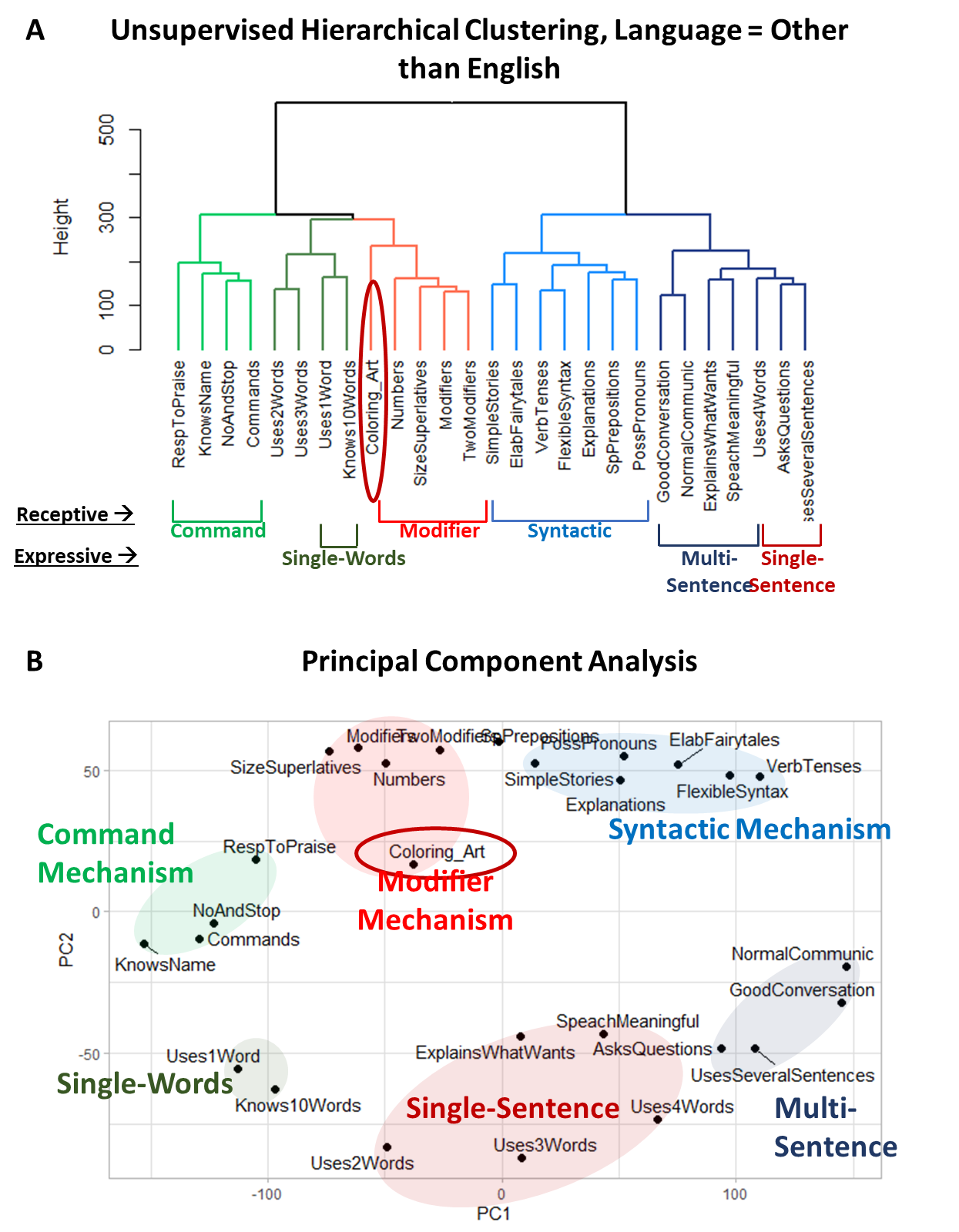
Supplementary Figure 11. Clustering analysis of 15 receptive language and 11 expressive language items, along with the caregiver-reported item “[My child] does drawing, coloring, art” (labeled *Coloring_Art*) in participants speaking languages other than English. (A) Dendrogram generated using UHCA. (B) PCA, with Principal Component 1 accounting for 33.3% of the variance in the data and Principal Component 2 accounting for 11.1%.

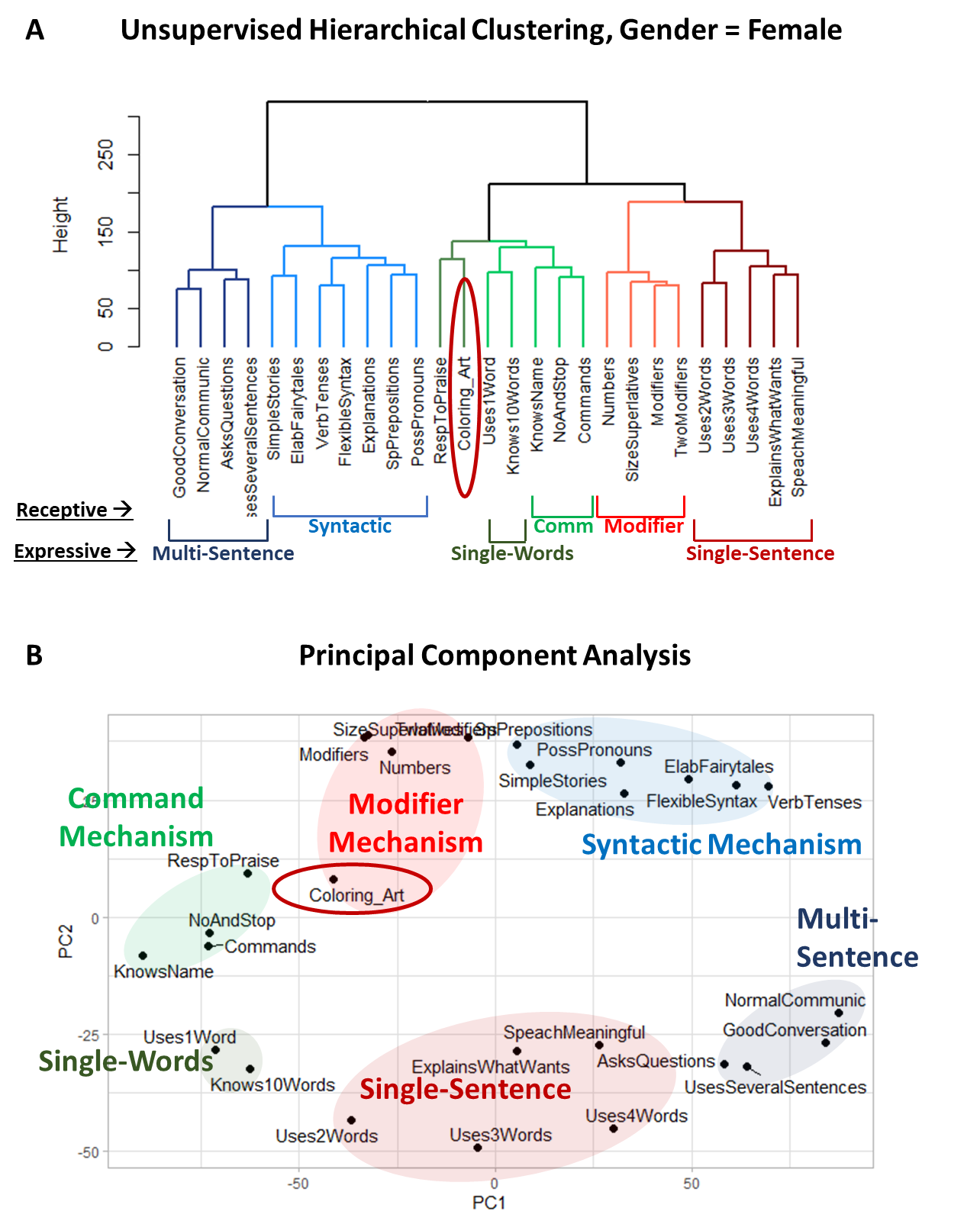
Supplementary Figure 12. Clustering analysis of 15 receptive language and 11 expressive language items, along with the caregiver-reported item “[My child] does drawing, coloring, art” (labeled *Coloring_Art*) in female participants. (A) Dendrogram generated using UHCA. (B) PCA, with Principal Component 1 accounting for 33.7% of the variance in the data and Principal Component 2 accounting for 11.5%.

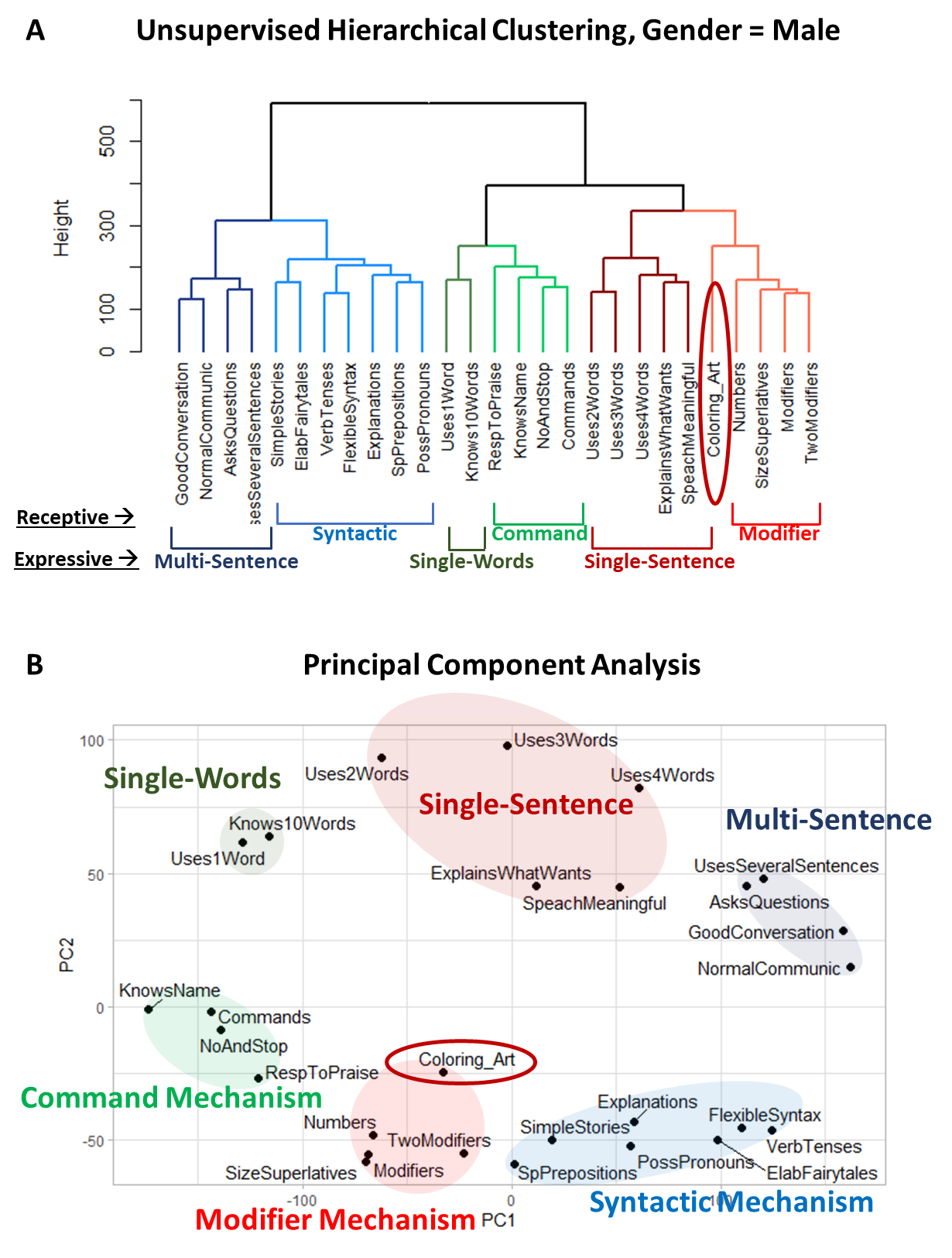
Supplementary Figure 13. Clustering analysis of 15 receptive language and 11 expressive language items, along with the caregiver-reported item “[My child] does drawing, coloring, art” (labeled *Coloring_Art*) in male participants. (A) Dendrogram generated using UHCA. (B) PCA, with Principal Component 1 accounting for 37.2% of the variance in the data and Principal Component 2 accounting for 10.3%.

### Clustering analysis of the item “[My child] draws a VARIETY of RECOGNIZABLE images (objects, people, animals, etc.)”

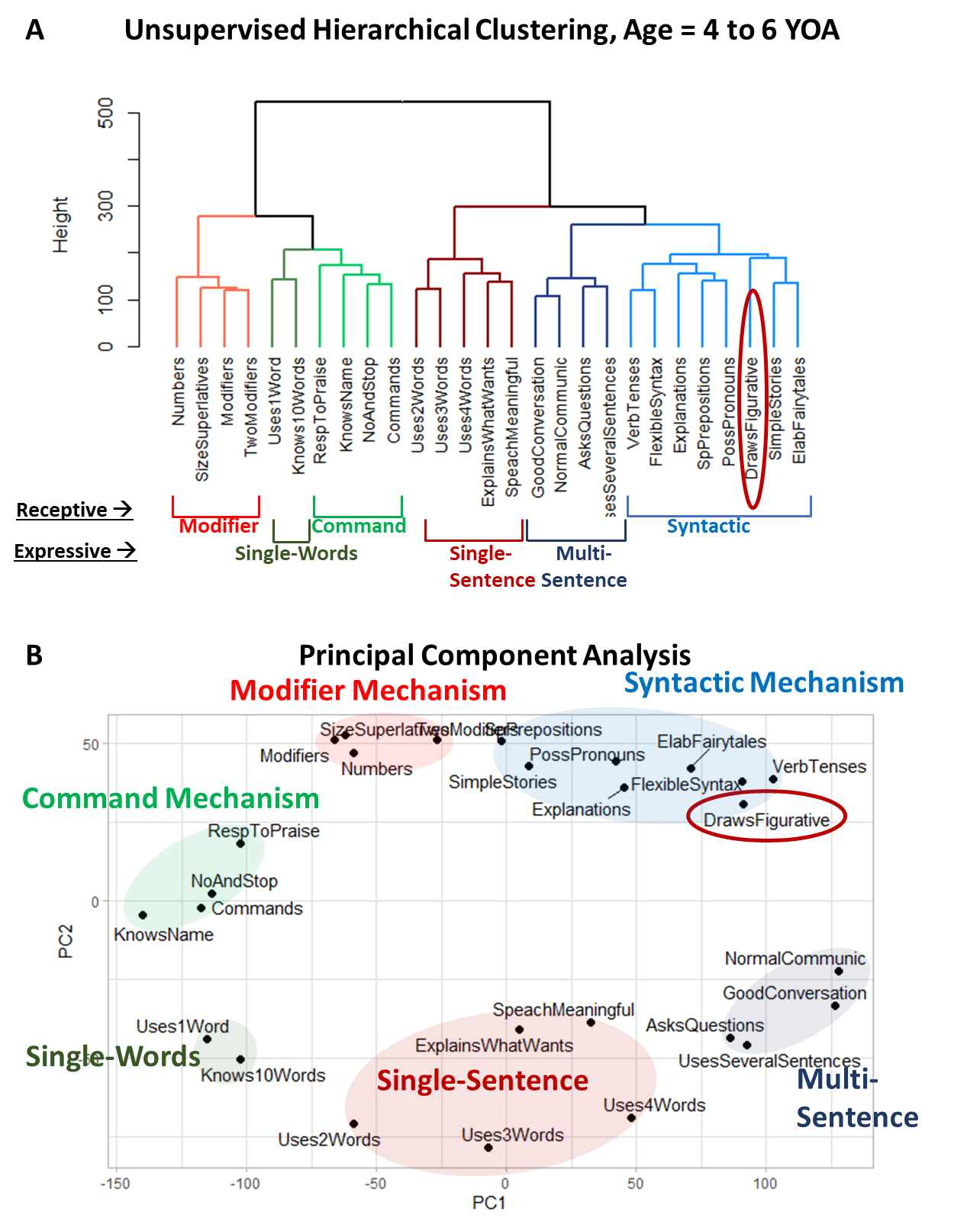
Supplementary Figure 14. Clustering analysis of 15 receptive language and 11 expressive language items along with the caregiver-reported item “[My child] draws a VARIETY of RECOGNIZABLE images (objects, people, animals, etc.)” labeled as *DrawsFigurative* in participants aged 4 to 6 years. (A) Dendrogram generated using UHCA. (B) PCA, with Principal Component 1 accounting for 36.5% of the variance in the data and Principal Component 2 accounting for 10.5%.

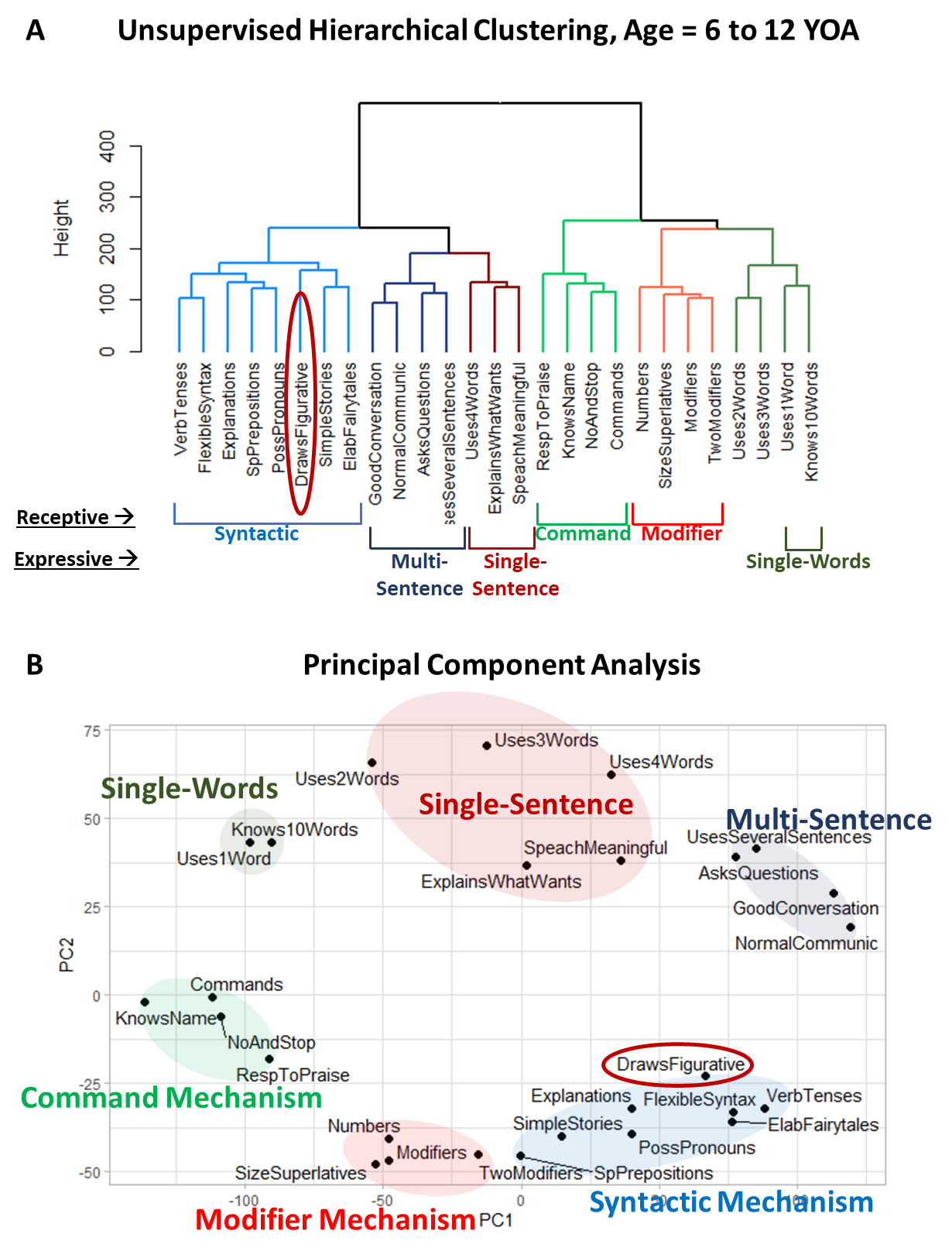
Supplementary Figure 15. Clustering analysis of 15 receptive language and 11 expressive language items along with the caregiver-reported item “[My child] draws a VARIETY of RECOGNIZABLE images (objects, people, animals, etc.)” labeled as *DrawsFigurative* in participants aged 6 to 12 years. (A) Dendrogram generated using UHCA. (B) PCA, with Principal Component 1 accounting for 37.5% of the variance in the data and Principal Component 2 accounting for 10.8%.

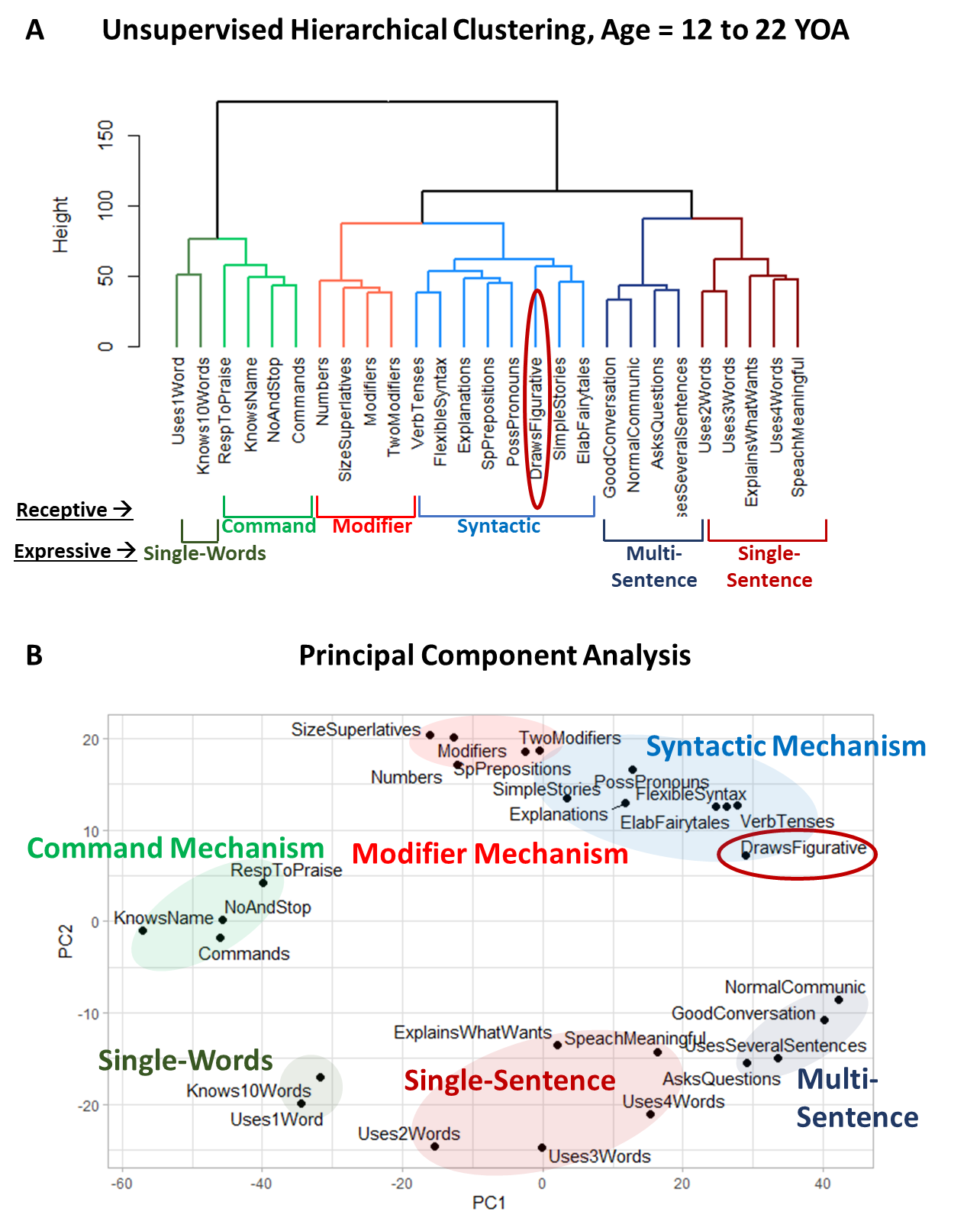
Supplementary Figure 16. Clustering analysis of 15 receptive language and 11 expressive language items along with the caregiver-reported item “[My child] draws a VARIETY of RECOGNIZABLE images (objects, people, animals, etc.)” labeled as *DrawsFigurative* in participants aged 12 to 22 years. (A) Dendrogram generated using UHCA. (B) PCA, with Principal Component 1 accounting for 38% of the variance in the data and Principal Component 2 accounting for 11.4%.

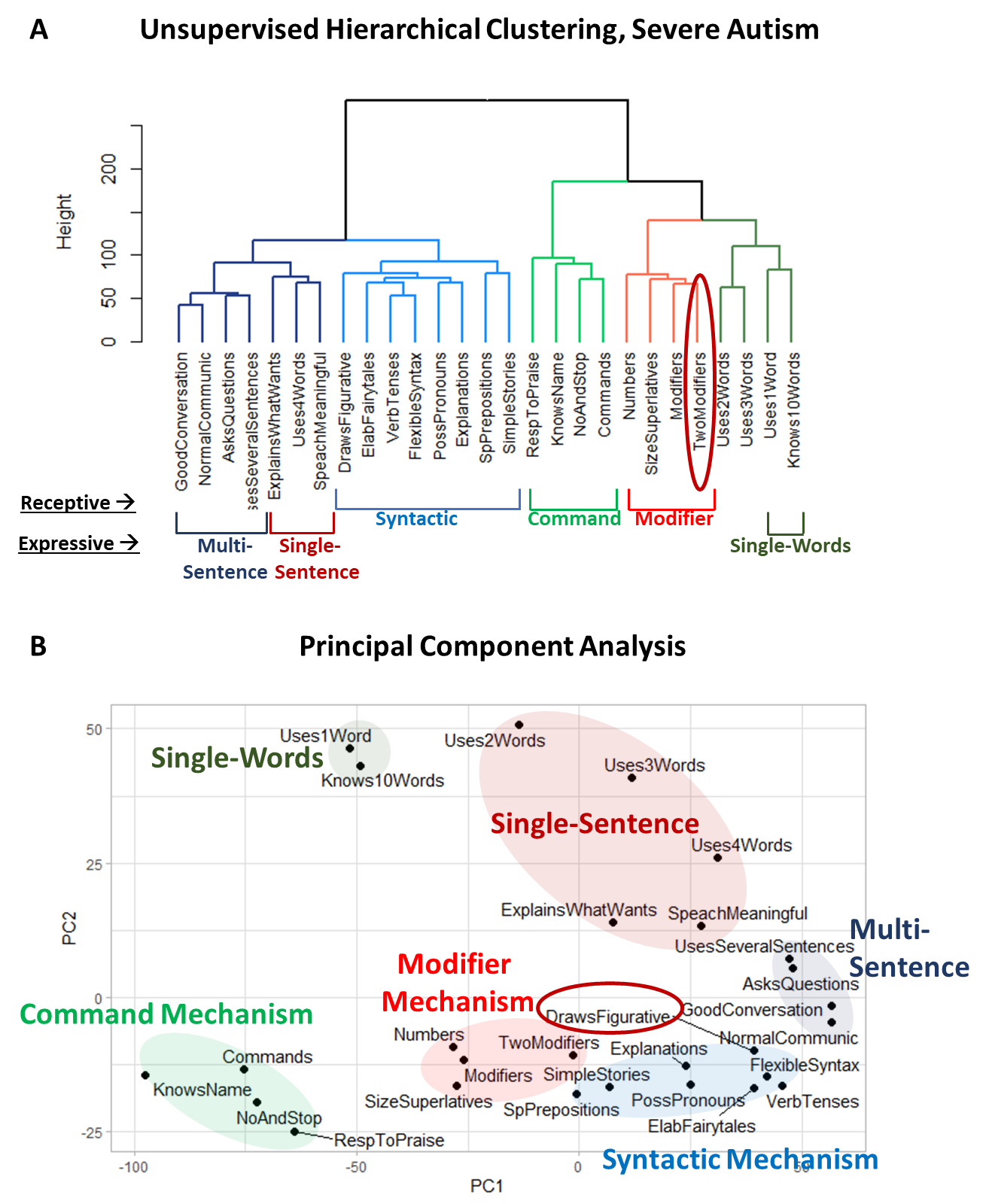
Supplementary Figure 17. Clustering analysis of 15 receptive language and 11 expressive language items along with the caregiver-reported item “[My child] draws a VARIETY of RECOGNIZABLE images (objects, people, animals, etc.)” labeled as *DrawsFigurative* in participants diagnosed with severe autism. (A) Dendrogram generated using UHCA. (B) PCA, with Principal Component 1 accounting for 39.3% of the variance in the data and Principal Component 2 accounting for 9.8%.

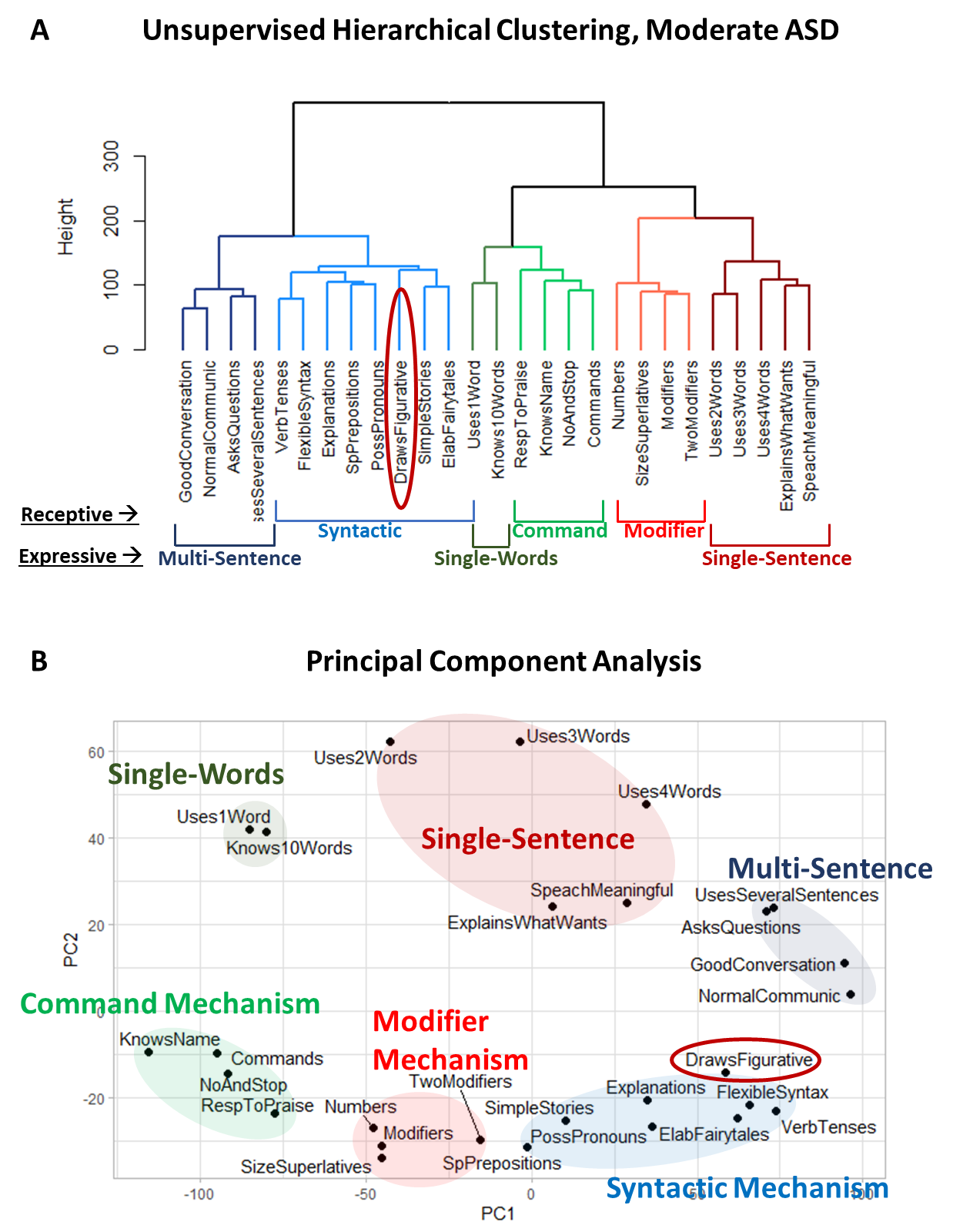
Supplementary Figure 18. Clustering analysis of 15 receptive language and 11 expressive language items along with the caregiver-reported item “[My child] draws a VARIETY of RECOGNIZABLE images (objects, people, animals, etc.)” labeled as *DrawsFigurative* in participants diagnosed with moderate autism. (A) Dendrogram generated using UHCA. (B) PCA, with Principal Component 1 accounting for 41.6% of the variance in the data and Principal Component 2 accounting for 9.6%.

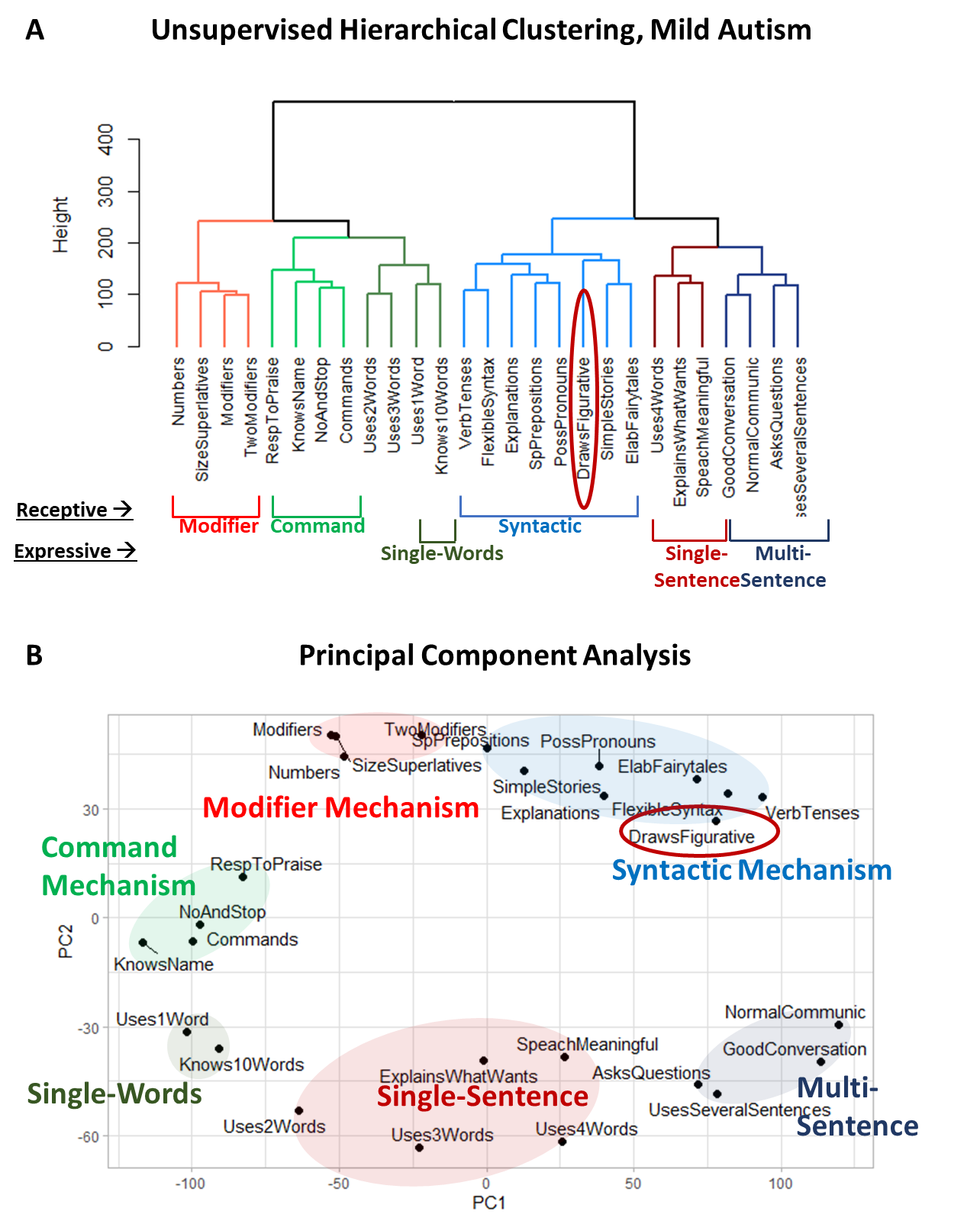
Supplementary Figure 19. Clustering analysis of 15 receptive language and 11 expressive language items along with the caregiver-reported item “[My child] draws a VARIETY of RECOGNIZABLE images (objects, people, animals, etc.)” labeled as *DrawsFigurative* in participants diagnosed with mild autism. (A) Dendrogram generated using UHCA. (B) PCA, with Principal Component 1 accounting for 36.4% of the variance in the data and Principal Component 2 accounting for 11.3%.

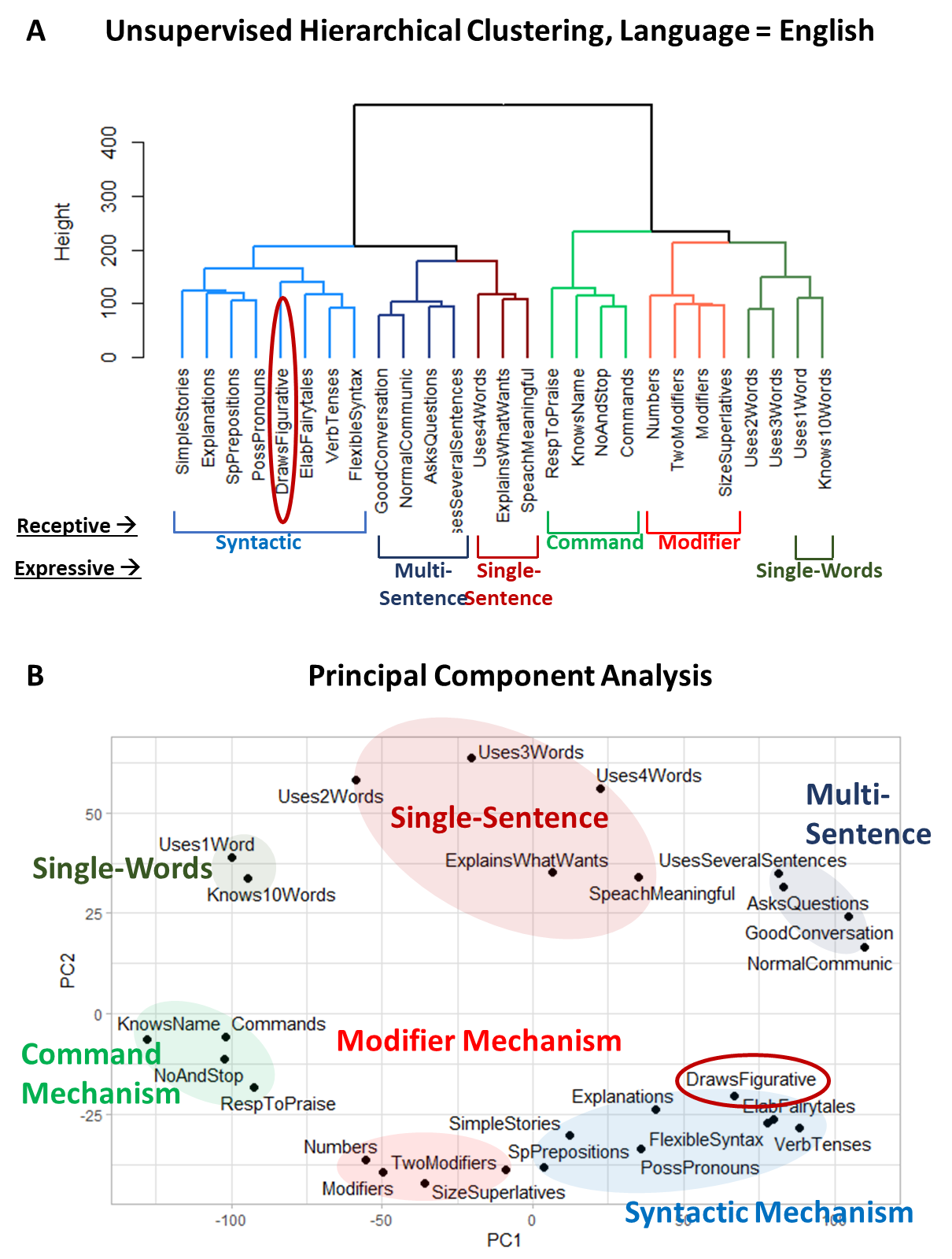
Supplementary Figure 20. Clustering analysis of 15 receptive language and 11 expressive language items along with the caregiver-reported item “[My child] draws a VARIETY of RECOGNIZABLE images (objects, people, animals, etc.)” labeled as *DrawsFigurative* in English-speaking participants. (A) Dendrogram generated using UHCA. (B) PCA, with Principal Component 1 accounting for 42.5% of the variance in the data and Principal Component 2 accounting for 9.6%.

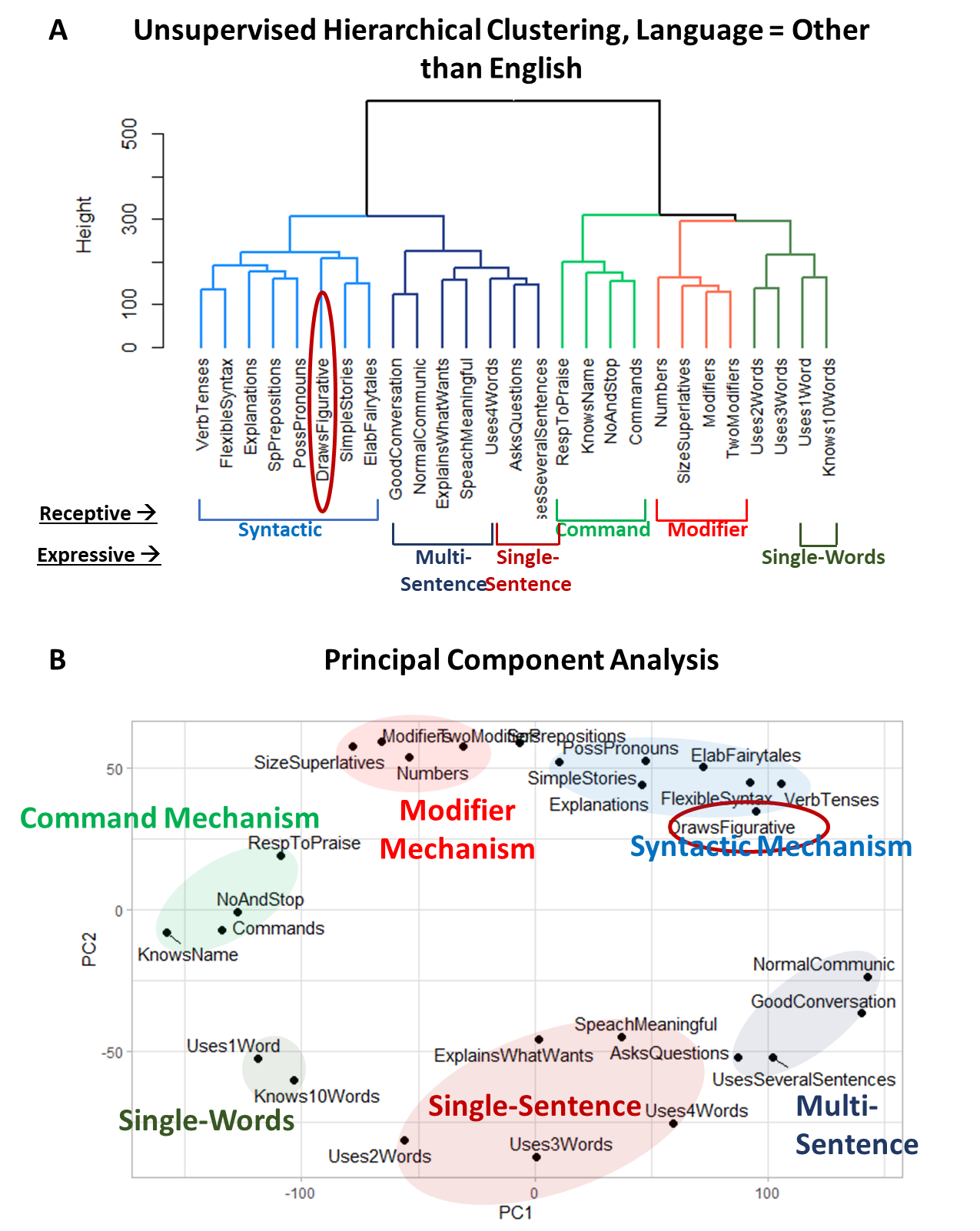
Supplementary Figure 21. Clustering analysis of 15 receptive language and 11 expressive language items along with the caregiver-reported item “[My child] draws a VARIETY of RECOGNIZABLE images (objects, people, animals, etc.)” labeled as *DrawsFigurative* in participants speaking languages other than English. (A) Dendrogram generated using UHCA. (B) PCA, with Principal Component 1 accounting for 34.2% of the variance in the data and Principal Component 2 accounting for 11.1%.

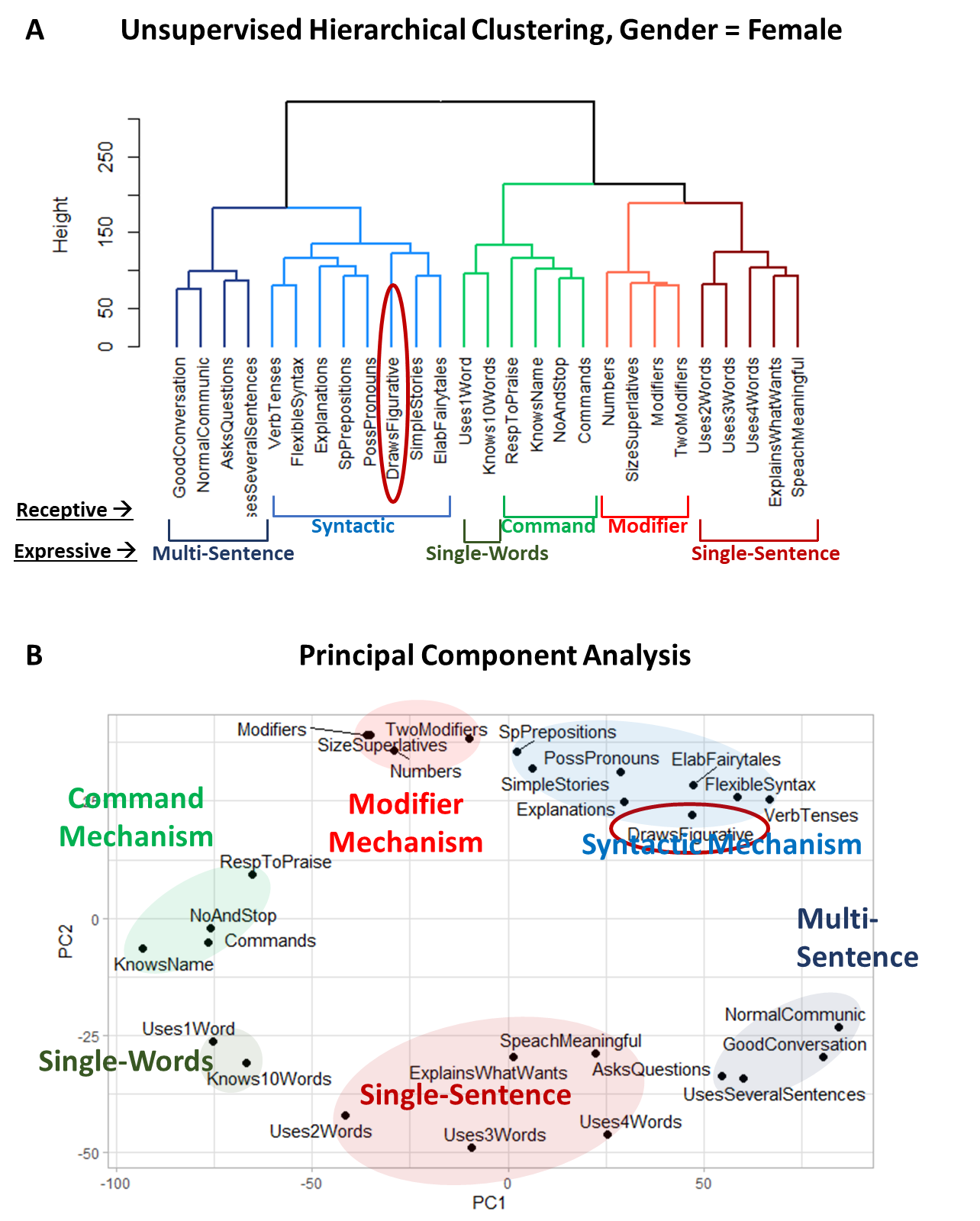
Supplementary Figure 22. Clustering analysis of 15 receptive language and 11 expressive language items along with the caregiver-reported item “[My child] draws a VARIETY of RECOGNIZABLE images (objects, people, animals, etc.)” labeled as *DrawsFigurative* in female participants. (A) Dendrogram generated using UHCA. (B) PCA, with Principal Component 1 accounting for 33.9% of the variance in the data and Principal Component 2 accounting for 11.6%.

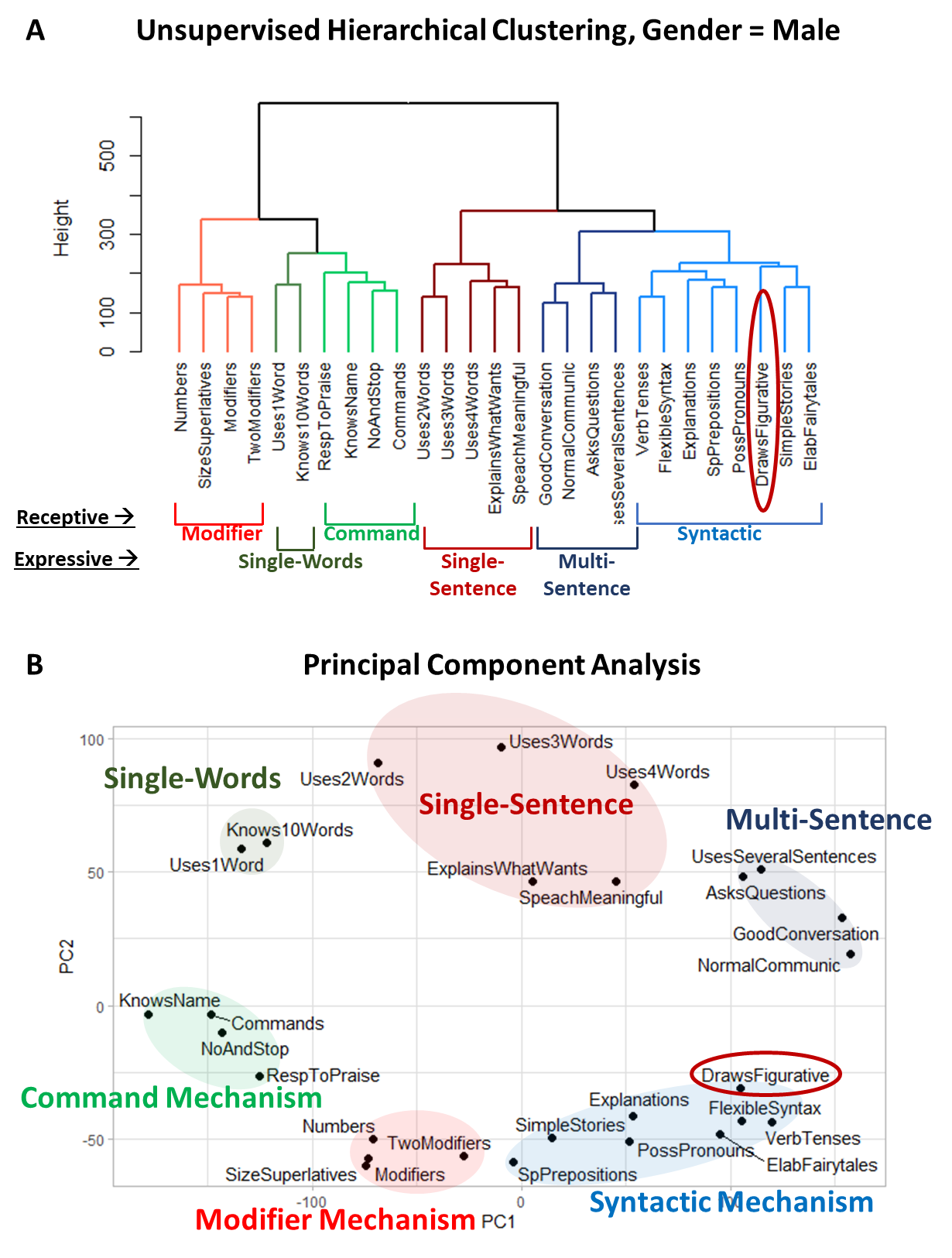
Supplementary Figure 23. Clustering analysis of 15 receptive language and 11 expressive language items along with the caregiver-reported item “[My child] draws a VARIETY of RECOGNIZABLE images (objects, people, animals, etc.)” labeled as *DrawsFigurative* in male participants. (A) Dendrogram generated using UHCA. (B) PCA, with Principal Component 1 accounting for 38.2% of the variance in the data and Principal Component 2 accounting for 10.2%.

### Clustering analysis of the item “[My child] can draw a NOVEL image following YOUR description (e.g. a three-headed horse)”

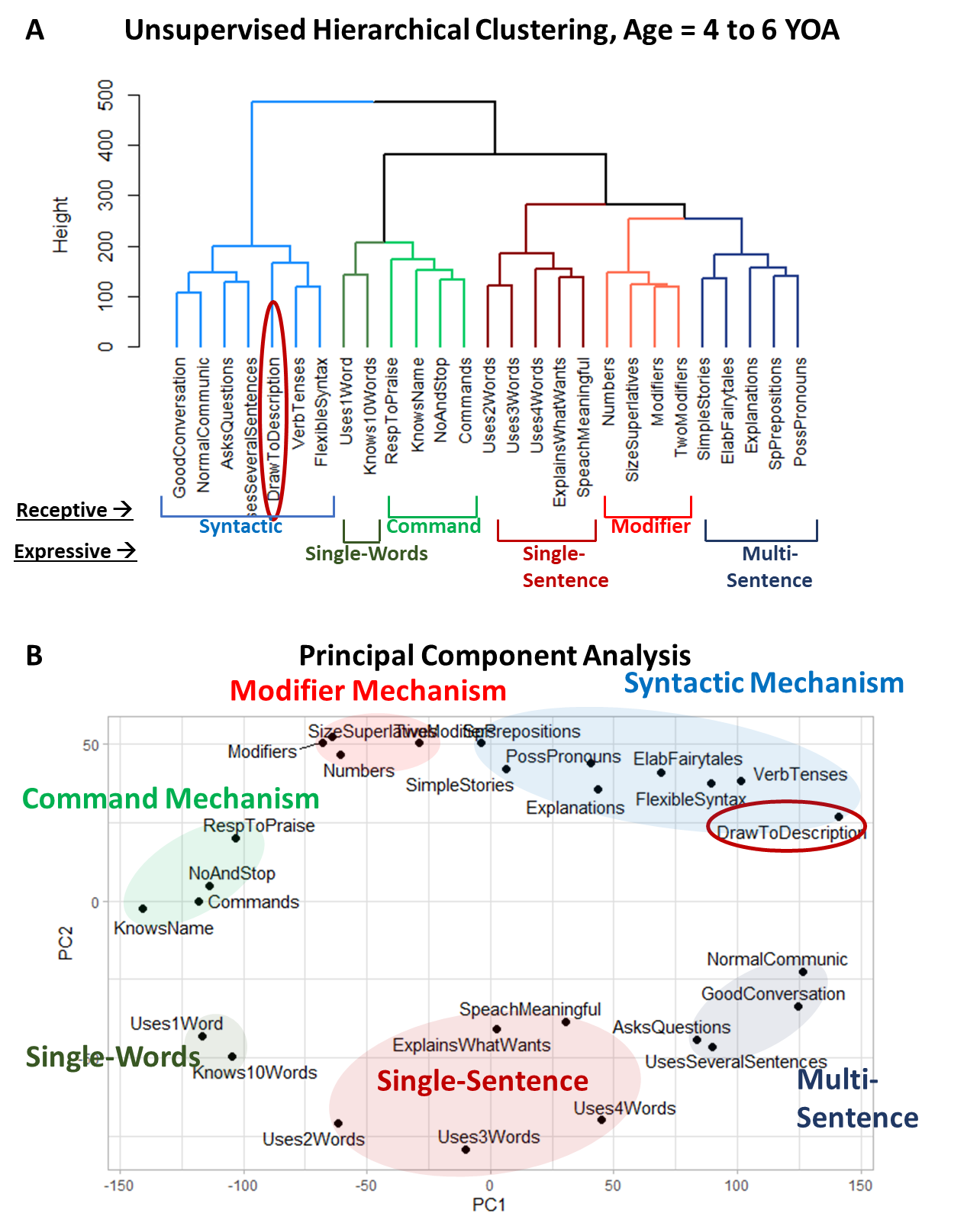
Supplementary Figure 24. Clustering analysis of 15 receptive language and 11 expressive language items along with the caregiver-reported item “[My child] draws a VARIETY of RECOGNIZABLE images (objects, people, animals, etc.)” labeled *DrawToDescritption* in participants aged 4 to 6 years. (A) Dendrogram generated using UHCA. (B) PCA, with Principal Component 1 accounting for 38.1% of the variance in the data and Principal Component 2 accounting for 10.3%.

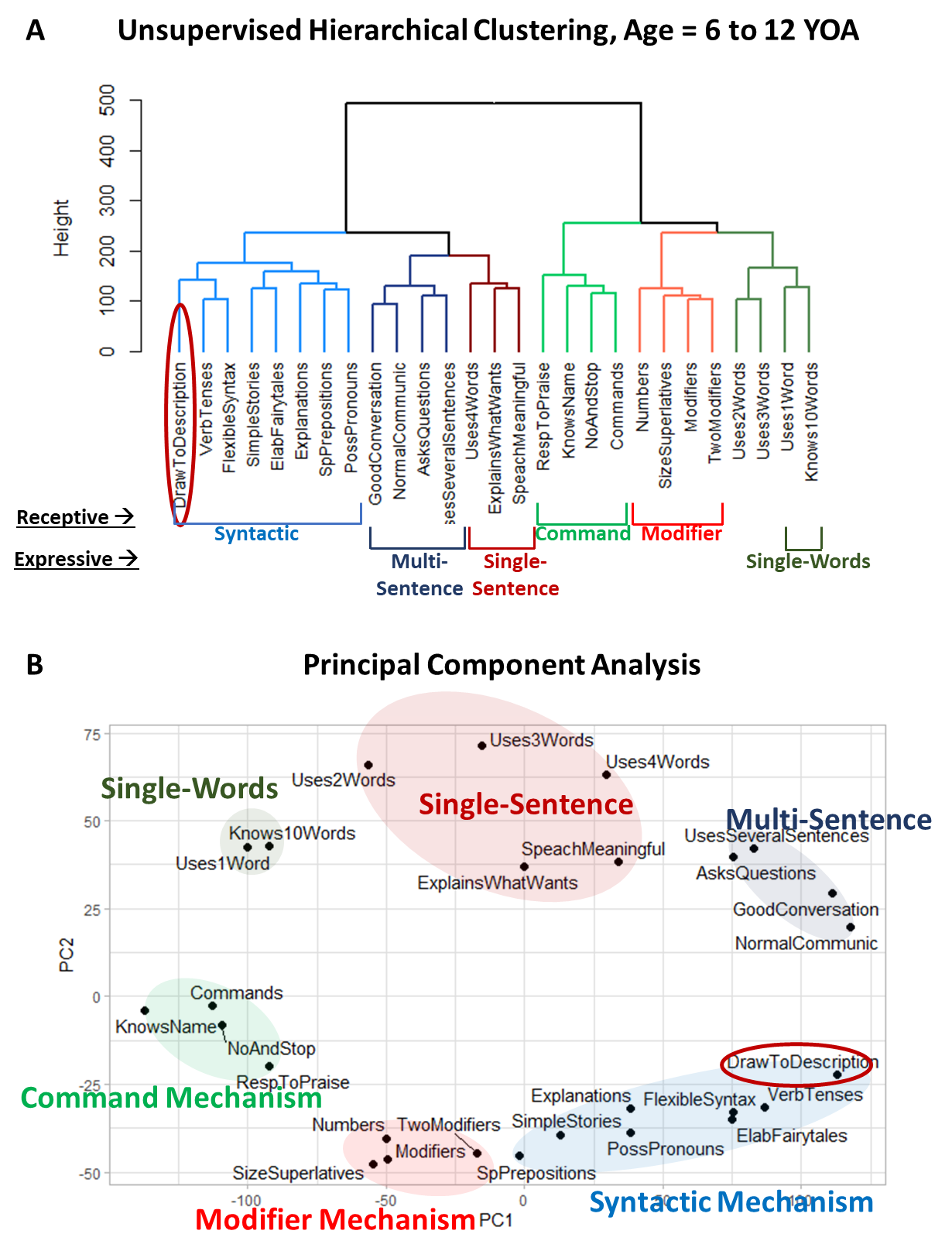
Supplementary Figure 25. Clustering analysis of 15 receptive language and 11 expressive language items along with the caregiver-reported item “[My child] draws a VARIETY of RECOGNIZABLE images (objects, people, animals, etc.)” labeled *DrawToDescritption* in participants aged 6 to 12 years. (A) Dendrogram generated using UHCA. (B) PCA, with Principal Component 1 accounting for 39.1% of the variance in the data and Principal Component 2 accounting for 10.7%.

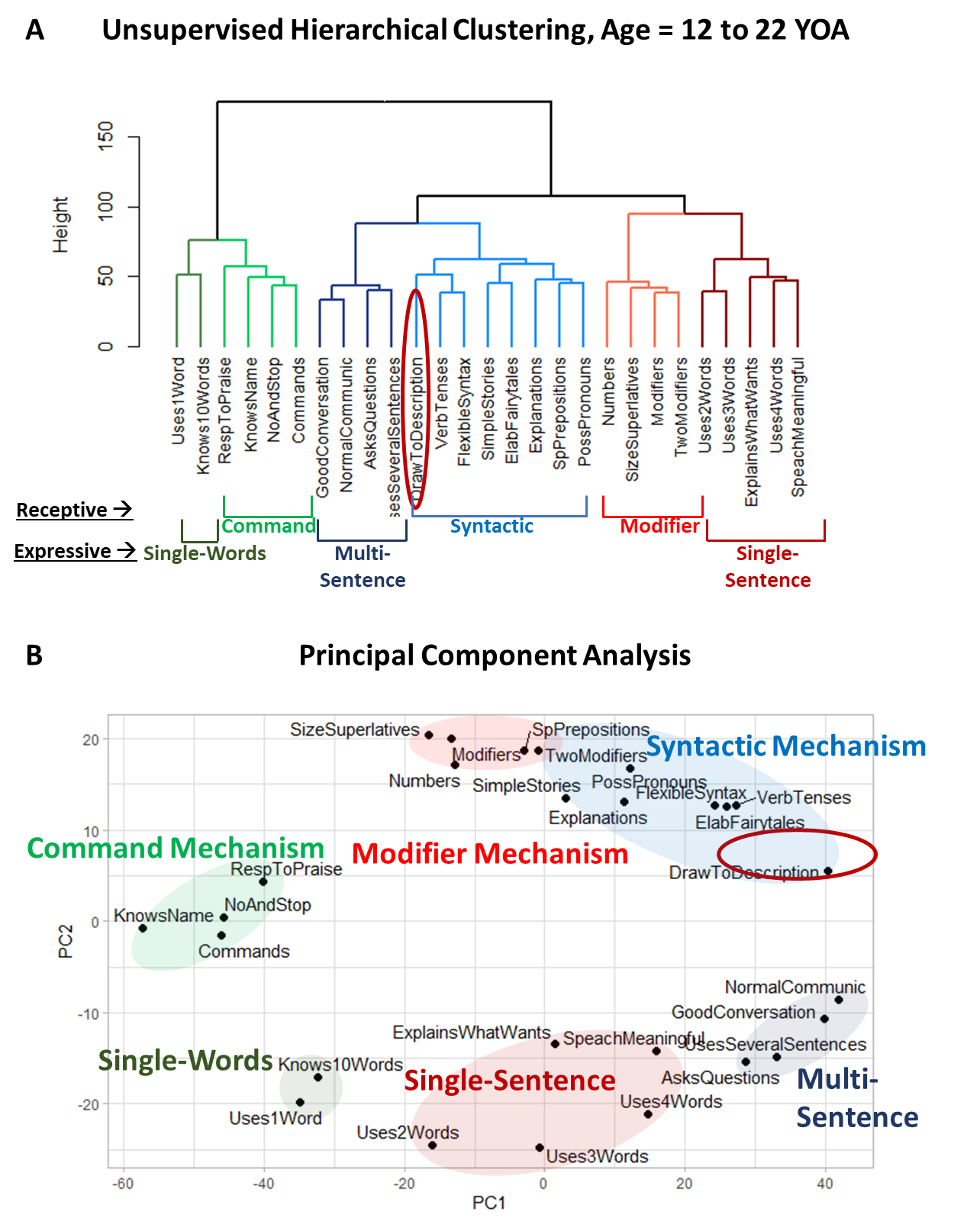
Supplementary Figure 26. Clustering analysis of 15 receptive language and 11 expressive language items along with the caregiver-reported item “[My child] draws a VARIETY of RECOGNIZABLE images (objects, people, animals, etc.)” labeled *DrawToDescritption* in participants aged 12 to 22 years. (A) Dendrogram generated using UHCA. (B) PCA, with Principal Component 1 accounting for 39.2% of the variance in the data and Principal Component 2 accounting for 11.3%.

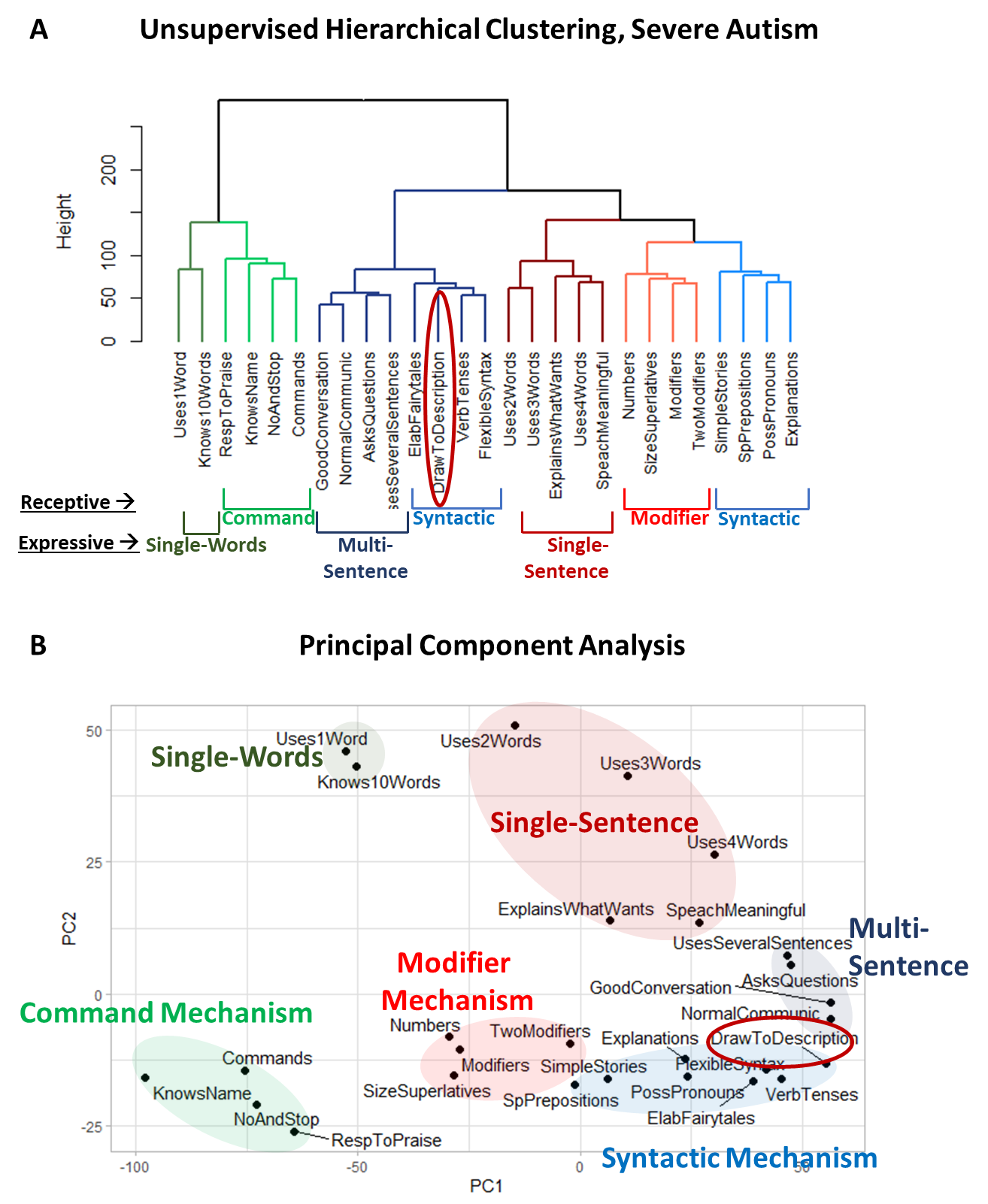
Supplementary Figure 27. Clustering analysis of 15 receptive language and 11 expressive language items along with the caregiver-reported item “[My child] draws a VARIETY of RECOGNIZABLE images (objects, people, animals, etc.)” labeled *DrawToDescritption* in participants diagnosed with severe autism. (A) Dendrogram generated using UHCA. (B) PCA, with Principal Component 1 accounting for 40.4% of the variance in the data and Principal Component 2 accounting for 9.9%.

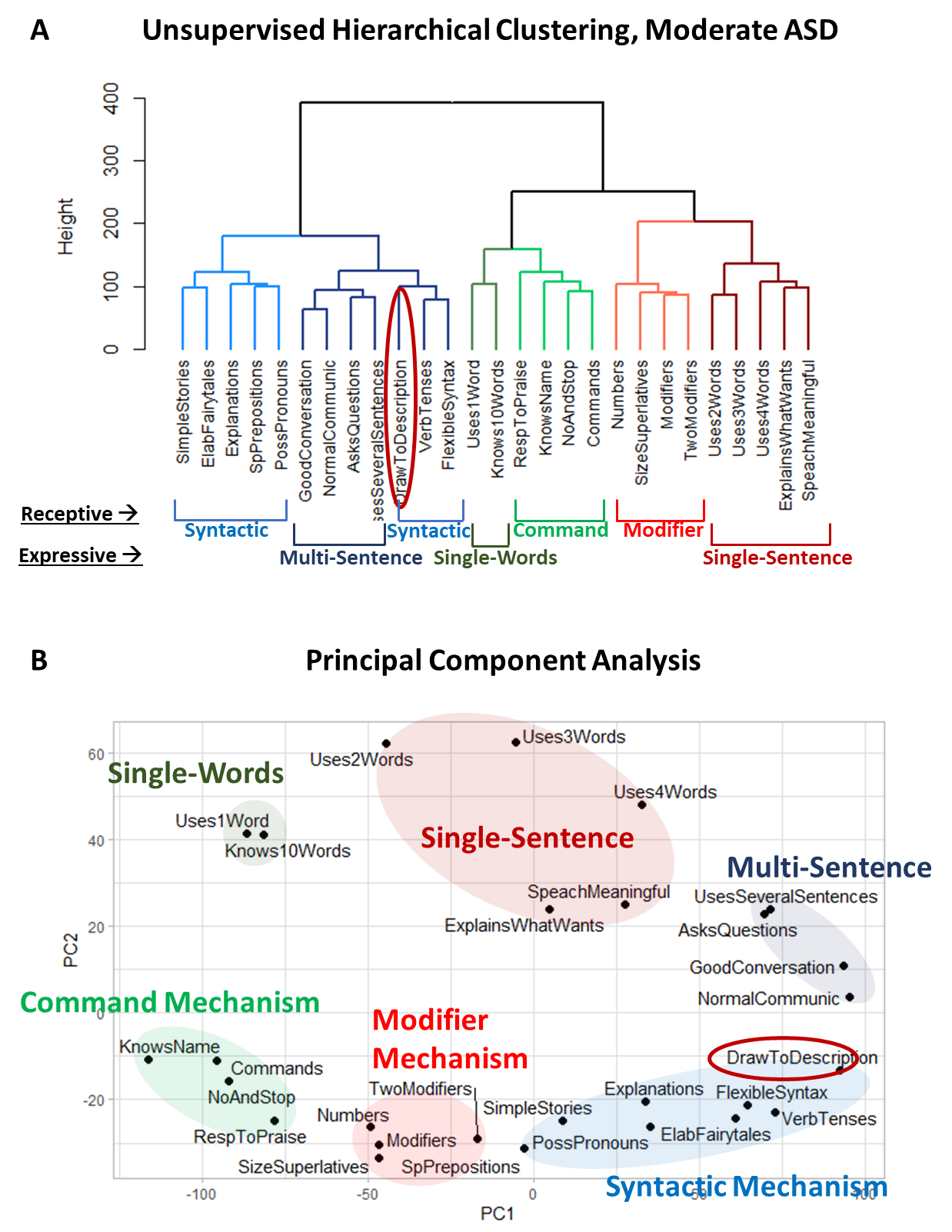
Supplementary Figure 28. Clustering analysis of 15 receptive language and 11 expressive language items along with the caregiver-reported item “[My child] draws a VARIETY of RECOGNIZABLE images (objects, people, animals, etc.)” labeled *DrawToDescritption* in participants diagnosed with moderate autism. (A) Dendrogram generated using UHCA. (B) PCA, with Principal Component 1 accounting for 43.2% of the variance in the data and Principal Component 2 accounting for 9.5%.

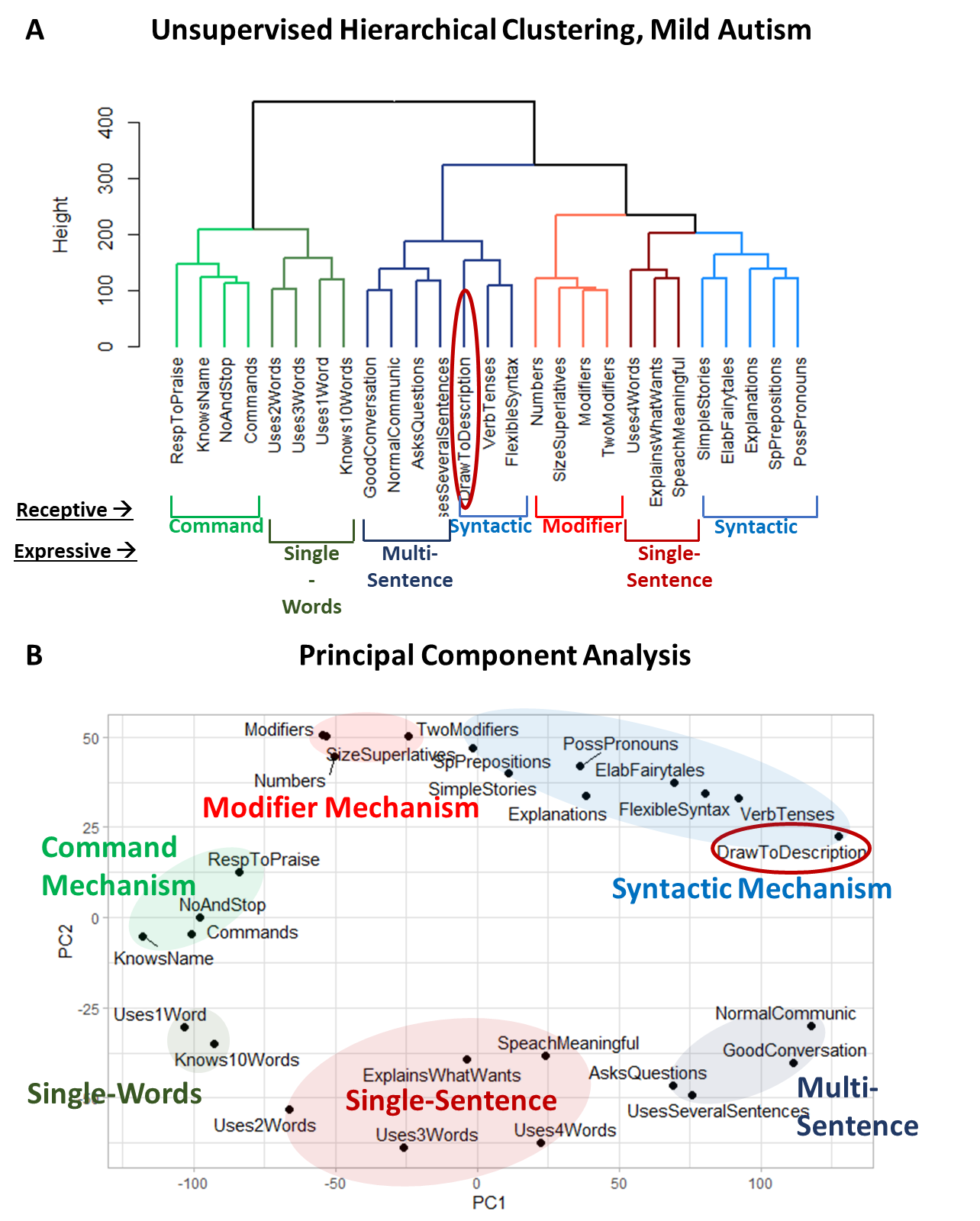
Supplementary Figure 29. Clustering analysis of 15 receptive language and 11 expressive language items along with the caregiver-reported item “[My child] draws a VARIETY of RECOGNIZABLE images (objects, people, animals, etc.)” labeled *DrawToDescritption* in participants diagnosed with mild autism. (A) Dendrogram generated using UHCA. (B) PCA, with Principal Component 1 accounting for 38.3 % of the variance in the data and Principal Component 2 accounting for 11.1%.

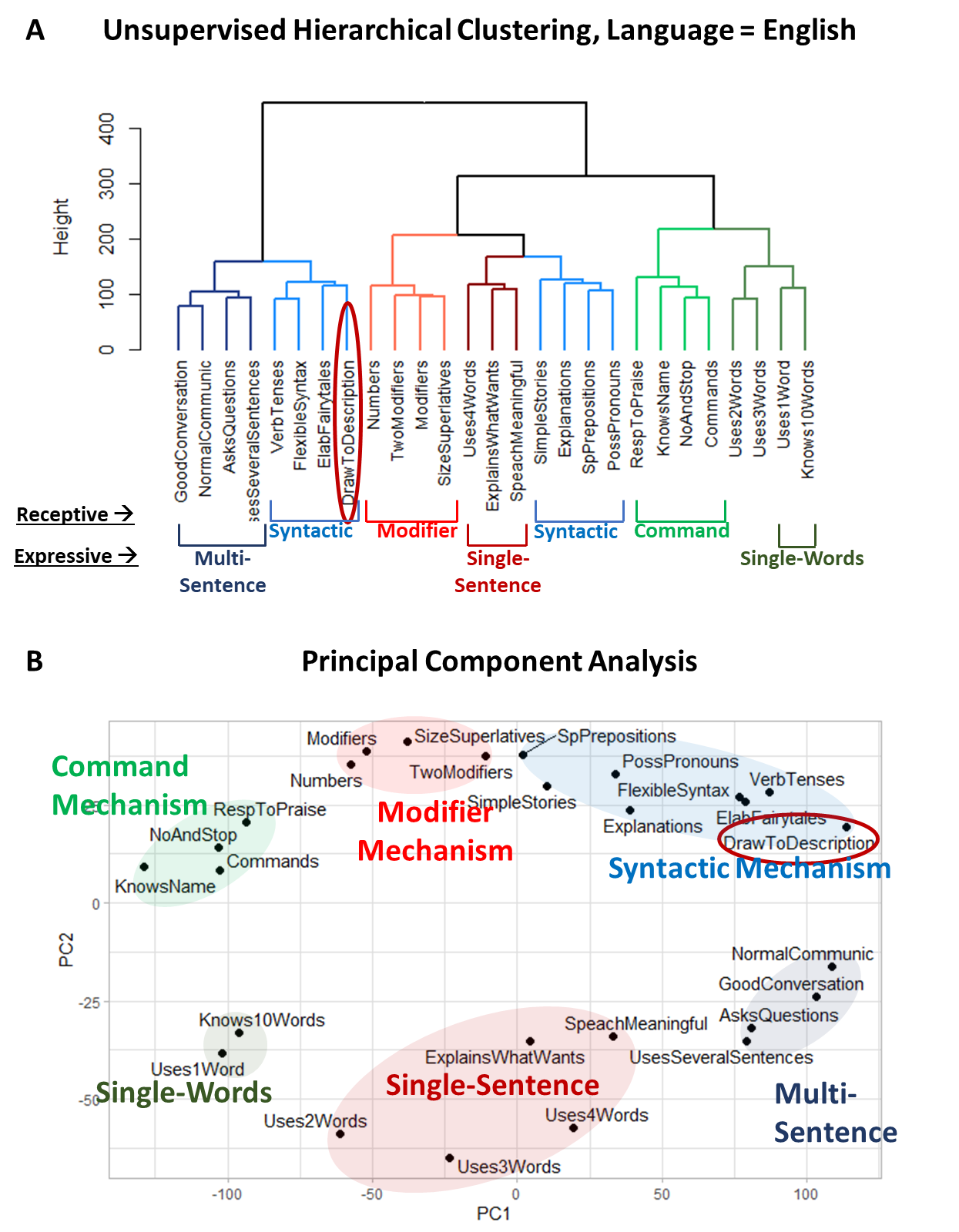
Supplementary Figure 29. Clustering analysis of 15 receptive language and 11 expressive language items along with the caregiver-reported item “[My child] draws a VARIETY of RECOGNIZABLE images (objects, people, animals, etc.)” labeled *DrawToDescritption* in English-speaking participants. (A) Dendrogram generated using UHCA. (B) PCA, with Principal Component 1 accounting for 44.4% of the variance in the data and Principal Component 2 accounting for 9.5%.

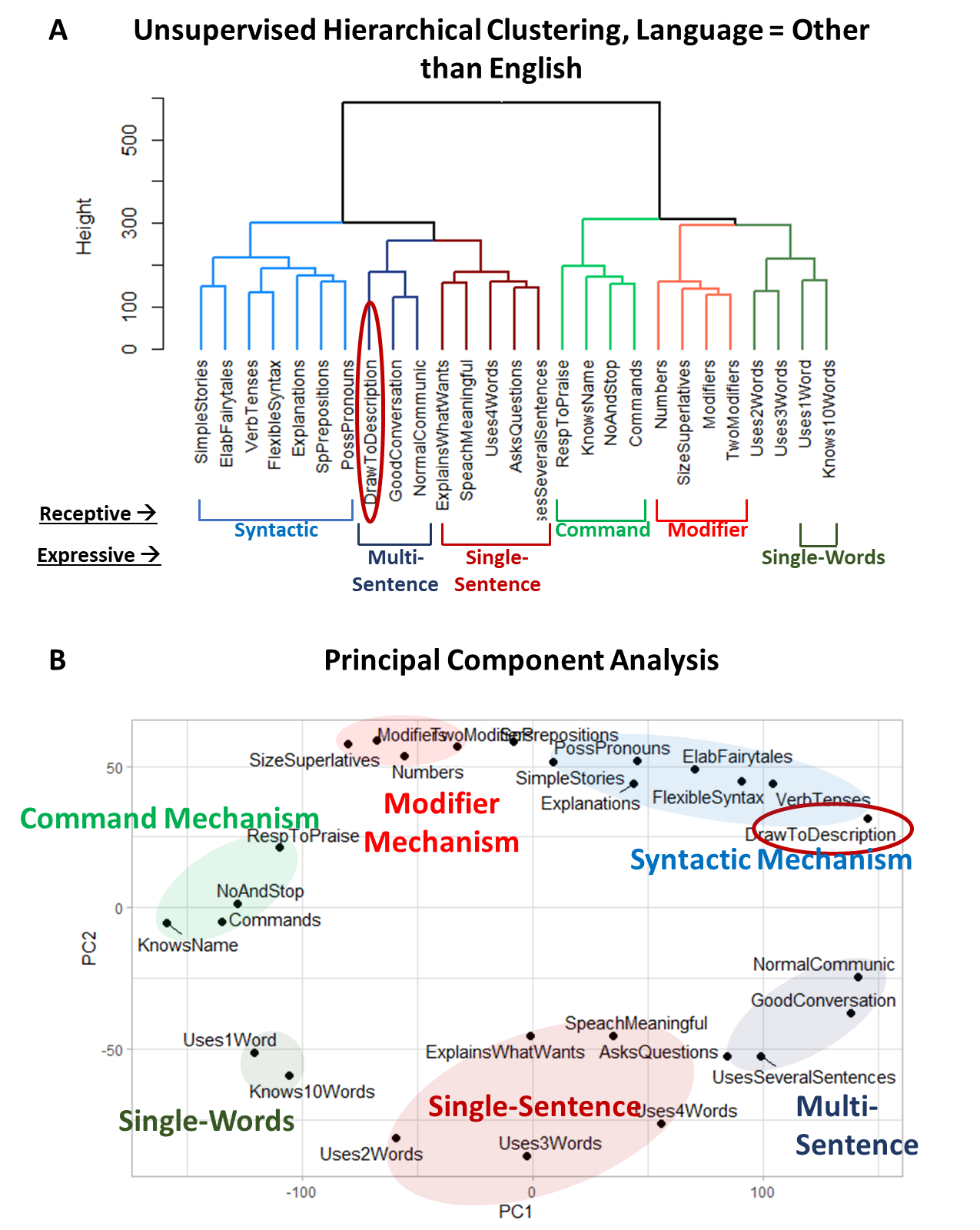
Supplementary Figure 30. Clustering analysis of 15 receptive language and 11 expressive language items along with the caregiver-reported item “[My child] draws a VARIETY of RECOGNIZABLE images (objects, people, animals, etc.)” labeled *DrawToDescritption* in participants speaking languages other than English. (A) Dendrogram generated using UHCA. (B) PCA, with Principal Component 1 accounting for 35.6% of the variance in the data and Principal Component 2 accounting for 11%.

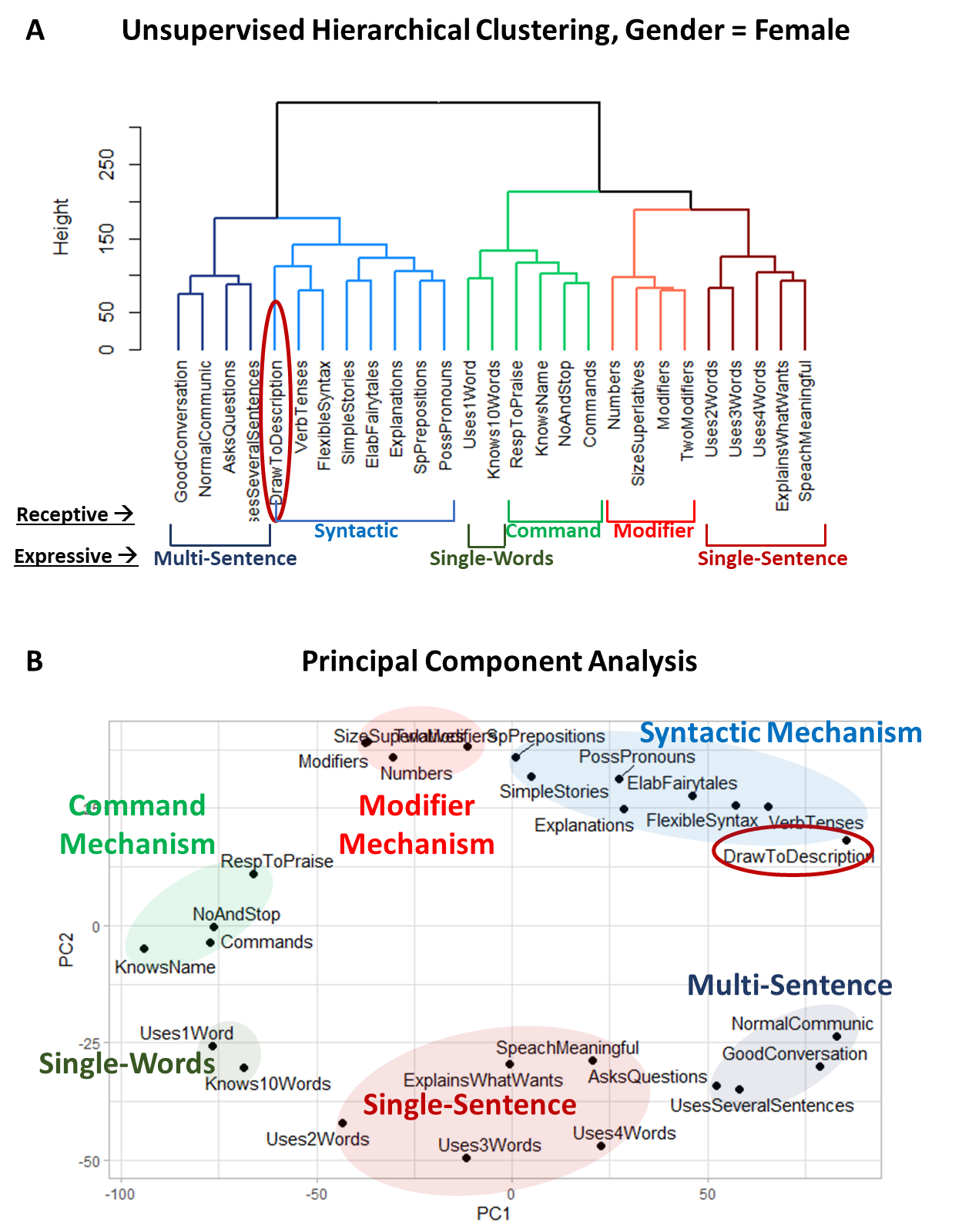
Supplementary Figure 31. Clustering analysis of 15 receptive language and 11 expressive language items along with the caregiver-reported item “[My child] draws a VARIETY of RECOGNIZABLE images (objects, people, animals, etc.)” labeled *DrawToDescritption* in female participants. (A) Dendrogram generated using UHCA. (B) PCA, with Principal Component 1 accounting for 35.5% of the variance in the data and Principal Component 2 accounting for 11.4%.

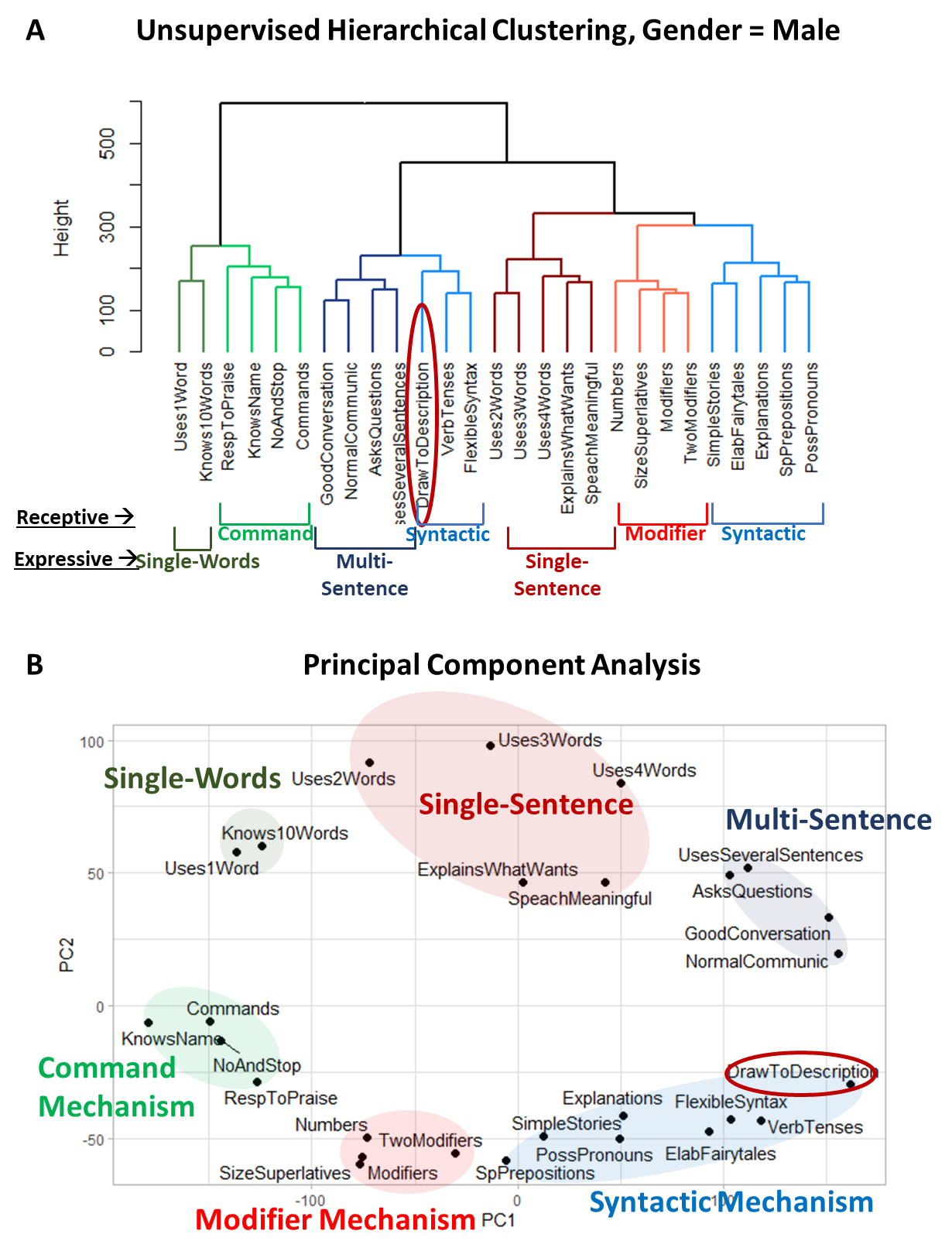
Supplementary Figure 31. Clustering analysis of 15 receptive language and 11 expressive language items along with the caregiver-reported item “[My child] draws a VARIETY of RECOGNIZABLE images (objects, people, animals, etc.)” labeled *DrawToDescritption* in male participants. (A) Dendrogram generated using UHCA. (B) PCA, with Principal Component 1 accounting for 39.8% of the variance in the data and Principal Component 2 accounting for 10.1%.
